# Evaluating the impacts of release in São Paulo State (Brazil) on the epidemic of covid-19 based on mathematical model

**DOI:** 10.1101/2020.08.03.20167221

**Authors:** Hyun Mo Yang, Luis Pedro Lombardi Junior, Ariana Campos Yang, Fabio Fernandes Morato Castro

## Abstract

To flatten the curve of the natural epidemic of covid-19, many countries adopted lockdown or isolation resulting in the containment of the SARS-CoV-2 transmission. However, an important question arises about the strategies of release of isolated persons to avoid overloaded hospitals and increased deaths. São Paulo State (Brazil) implemented the isolation of the population in non-essential activities on March 24, and the progressive flexibilization considering the characteristics of each location (release of the isolated population) initiated on June 15. A mathematical model based on the natural history of covid-19 was applied to describe the epidemiological scenario with isolation in São Paulo State, and assess the impact of release on the covid-19 epidemic. Using data collected from São Paulo State, we estimated the model parameters to obtain the curves of the epidemic, the number of deaths, and the clinical evolution of covid-19. The epidemic under isolation was the framework to evaluate the strategies of the release, that is, how these curves are changed with the release of isolated persons. We evaluated three strategies of release. First two strategies considered four releases in the isolated population in four equal proportions, but successive releases elapsed by 14 and 21 days. In each strategy the beginning of the release was on June 29 and July 13, when the effective reproduction number *R_ef_* was evaluated. The third strategy aimed at the protection of the elder subpopulation. We observed that the delay to begin the release and the increased elapse between successive releases resulted in a better scenario by decreasing severe covid-19 cases and, consequently, to avoid overloaded hospitals. We also observed that the release delayed to achieve lower values for *R_ef_* and infectious persons retarded in several months the quick increasing phase of the forthcoming epidemic. However, this epidemic can be flattened or even suppressed by isolation of infectious persons by mass testing and/or by rigid adoption of protective measures and social distancing.

## I. INTRODUCTION

The World Health Organization (WHO) declared coronavirus disease 2019 (covid-19) a pandemic on March 11, 2020. The severe acute respiratory syndrome coronavirus 2 (SARS-CoV-2) infects susceptible persons through the nose, mouth, or eyes, and infects cells in the respiratory tract. Shi et al. [1] divided the infection by SARS-CoV-2 into three stages: (1) asymptotic incubation period, (2) non-severe symptomatic period with detectable virus, and (3) severe respiratory symptomatic with high viral load. Covid-19 in mild form presents fever, dry cough, chills, malaise, muscle pain, and sore throat, in moderate form presents fever, respiratory symptoms, and radiographic characteristics, and in severe form manifests dyspnea, low oxygen saturation, and may evolve to respiratory failure, and multiple organ failure.

In the absence of effective treatment and vaccine, the control of the SARS-CoV-2 with high transmissibility and lethality many countries adopted lockdown at the very beginning of the epidemic. However, less rigid lockdown, known as isolation, can also be adopted, as did São Paulo State (Brazil). In this case, in the circulating population, the epidemic curve is flattened to avoid the overloading in hospitals, and the immunization by the infection is increased compared to the lockdown. Unfortunately, the number of deaths due to covid-19 increases. However, the isolated population must be released, which raises the question of a better strategy of release to avoid the collapse of the health care system. The deleterious effects of overloaded hospitals are possibly the death of untreated severe covid-19 cases and the stress of the health care workers besides an increased infection among them.

To incorporate the impact of SARS-CoV-2 transmission on the number of patients in hospitals, we evaluate the clinical evolution of severe covid-19. Siddiqi et al. [2] described three progressive phases of covid-19: (1) mild (early infection), (2) moderate, subdivided into without and with hypoxia (pulmonary involvement), and (3) severe (systemic hyper inflammation). The new cases of severe covid-19 are distributed into these three stages of disease progression to assess the number of patients needing hospitals and ICUs.

In [3], a mathematical model based on the natural history of covid-19 encompassing the different fatality rates in young (60 years old or less) and elder (60 years old or more) sub-populations was developed aiming to study the impacts of the isolation and further release on the epidemic of covid-19. That model was applied to São Paulo State to describe the epidemiological scenarios considering intermittent pulses in isolation and releases. That model was improved in [4] allowing the transmission of the infection by persons presenting mild covid-19 symptoms, and incorporating the protective measures that reduced the transmission of the virus. That model was applied to evaluate the impacts on the control of the covid-19 epidemic by isolation in São Paulo State and lockdown in Spain associated with the protective measures (washing hands with alcohol and gel, use of face mask, and social distancing).

In this paper, we apply the model presented in [4] to provide the epidemiological scenarios of release based on the plan elaborated by the São Paulo State authorities [5]. We observed that the epidemiological values (numbers of severe covid-19 cases, immune persons, and deaths) achieved in all strategies of release, except those maintaining elder persons isolated until the ending phase of the epidemic after the release of young persons, were practically those found in the natural epidemic (without any interventions). For this reason, the evaluation of the occupancy in hospitals and ICUs during the epidemic under release is important to avoid the overload of the health care system. However, the adoption of more rigorous protective measures and the isolation of infected persons caught by mass testing can decrease the epidemic under release.

## II. MATERIALS AND METHODS

In this section, we describe briefly the model developed in [4]. From the model, we propose a method to evaluate the clinical evolution of severe covid-19 and deaths.

### A. Mathematical model

A population is divided into two groups, composed of young (60 years old or less, denoted by subscript *y*), and elder (60 years old or more, denoted by subscript *o*) persons. The natural history of the new coronavirus infection is the same for young (*y*) and elder (*o*) subpopulations. Each subpopulation *j* (*j* = *y*, *o*) is divided into nine classes. The classes *S_j_* and *Q_j_* do not harbor virus, and class *E_j_* is composed of persons who had contact with the virus, but are not infectious. The classes *A_j_*, *D*_1_*_j_*, and a fraction *z_j_* of *Q*_2_*_j_* are composed of infectious persons, while classes *Q*_1_*_j_*, *D*_2_*_j_*, *Q*_3_*_j_*, and a fraction 1 − *z_j_* of *Q*_2_*_j_* are isolated and do not transmit the virus. Finally, class *I* is composed of immune persons. Table I summarizes the model classes (or variables).

**TABLE I:** Summary of the model variables (*j* = *y*, *o*).

| Symbol | Meaning |
| --- | --- |
| $S_j$ | Susceptible persons |
| $Q_j$ | Isolated among susceptible persons |
| $E_j$ | Exposed and incubating new coronavirus persons |
| $A_j$ | Asymptomatic persons |
| $D_{1j}$ | Pre-diseased (pre-symptomatic) persons |
| $Q_{1j}$ | Pre-diseased persons caught by test |
| $Q_{2j}$ | Mild (non-hospitalized) covid-19 persons |
| $Q_{3j}$ | Mild covid-19 persons adhered to isolation |
| $D_{2j}$ | Severe (hospitalized) covid-19 persons |
| $I$ | Immune (recovered) persons |

Susceptible persons are infected according to *λ_j_ S_j_* (known as the mass action law [6]) and enter into class *E_j_*, where *λ_j_* is the per-capita incidence rate (or force of infection) defined by *λ_j_* = *λ* (*δ_jy_* + *ψδ_jo_*), with *λ* being

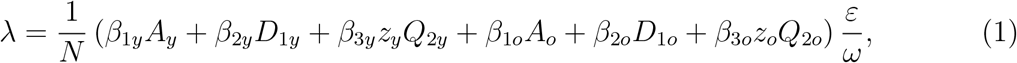

where *δ_ij_* is the Kronecker delta, with *δ_ij_* = 1 if *i* = *j*, and 0 if *i* ≠ *j*; and *β*_1_*_j_*, *β*_2_*_j_* and *β*_3_*_j_* are the transmission rates, that is, the rates at which a virus encounters a susceptible people and infects him/her. These transmission rates depend on the social behavior (network of contacts), demographic density, interaction between infectious and susceptible persons, and the transmissibility of the virus. The protection factor *ε* (*ε* ≤ 1) decreases the transmission of infection by individual (face mask, hygiene, etc.) and collective (social distancing) protective measures, while the reduction factor *ω* (*ω* ≥ 1) decreases the transmission rates (populational density and social contacts diminish in small cities). Table II summarizes the model parameters described in Appendix A.

**TABLE II:**
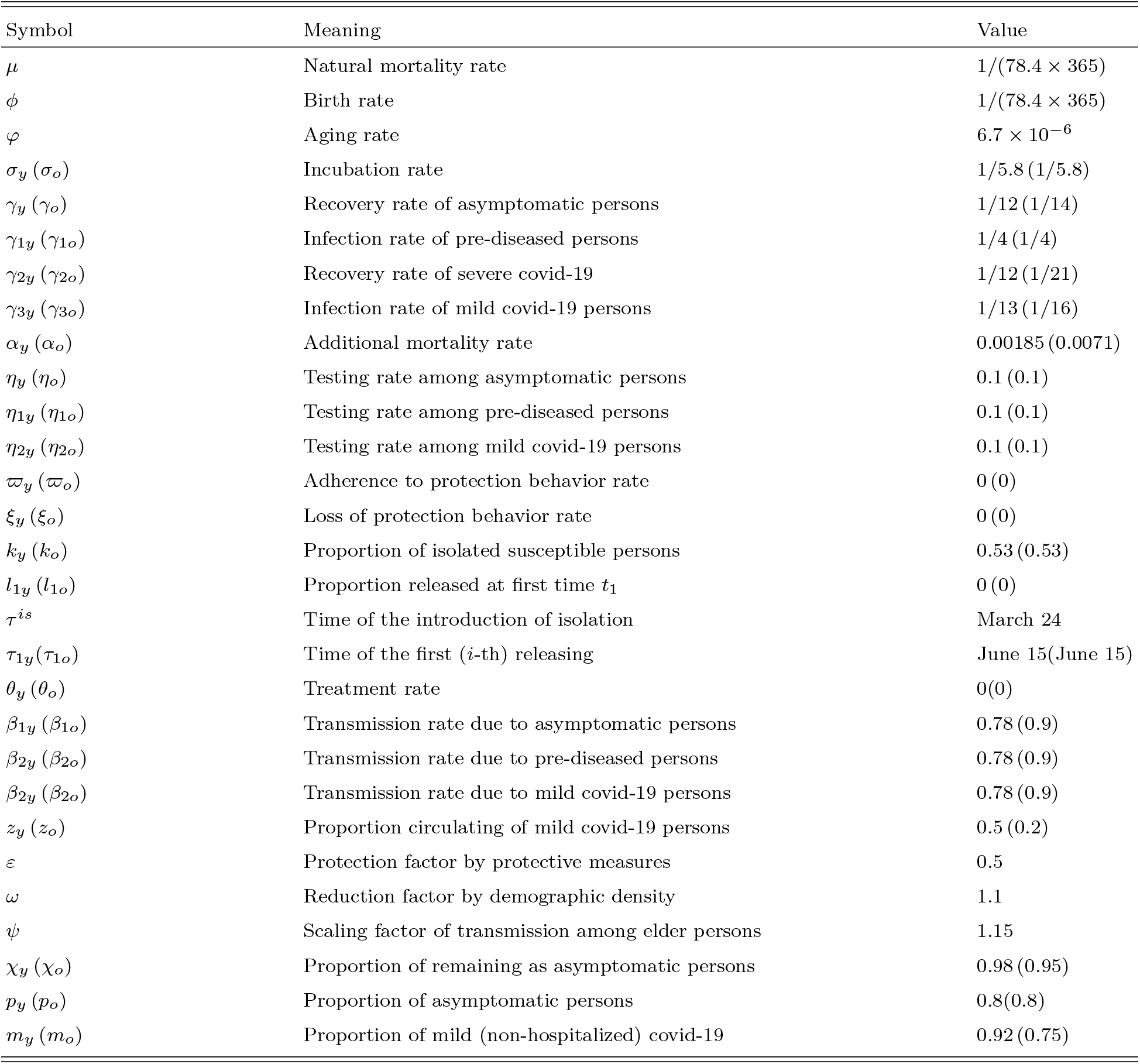
Summary of the model parameters (*j* = *y*, *o*) and values (rates in days ^−1^, time in days and proportions are dimensionless).

To incorporate the isolation and further release, we consider the following equations for those who did not have contact with the virus

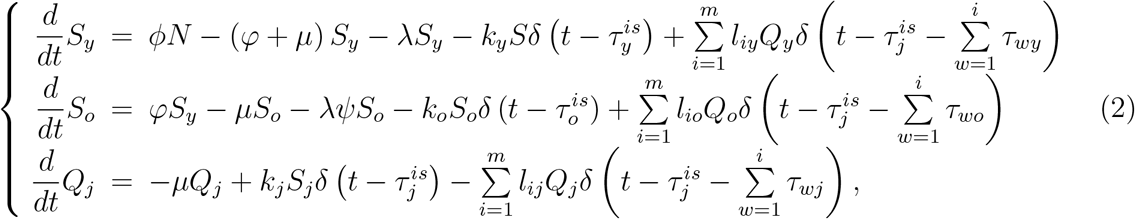

where a unique pulse in isolation is adopted at time 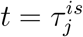, described by 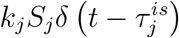, and *m* intermittent releases are described by 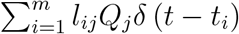, where 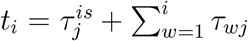 for *j* = *y*, *o*, and *δ* (*x*) is the Dirac delta function, that is, *δ* (*x*) = *∞*, if *x* = 0, otherwise, *δ* (*x*) = 0 with 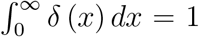. The fraction of persons in isolation is *k_j_*, and /*l_ij_*, *i* = 1, 2, ···, *m*, is the fraction of the *i*-th release of isolated persons, with *τ_wj_* being the period between successive releases. For other variables, the dynamic equations are obtained through the balance between inflow and outflow as shown in Figure 19 in Appendix A.

**FIG. 19:**
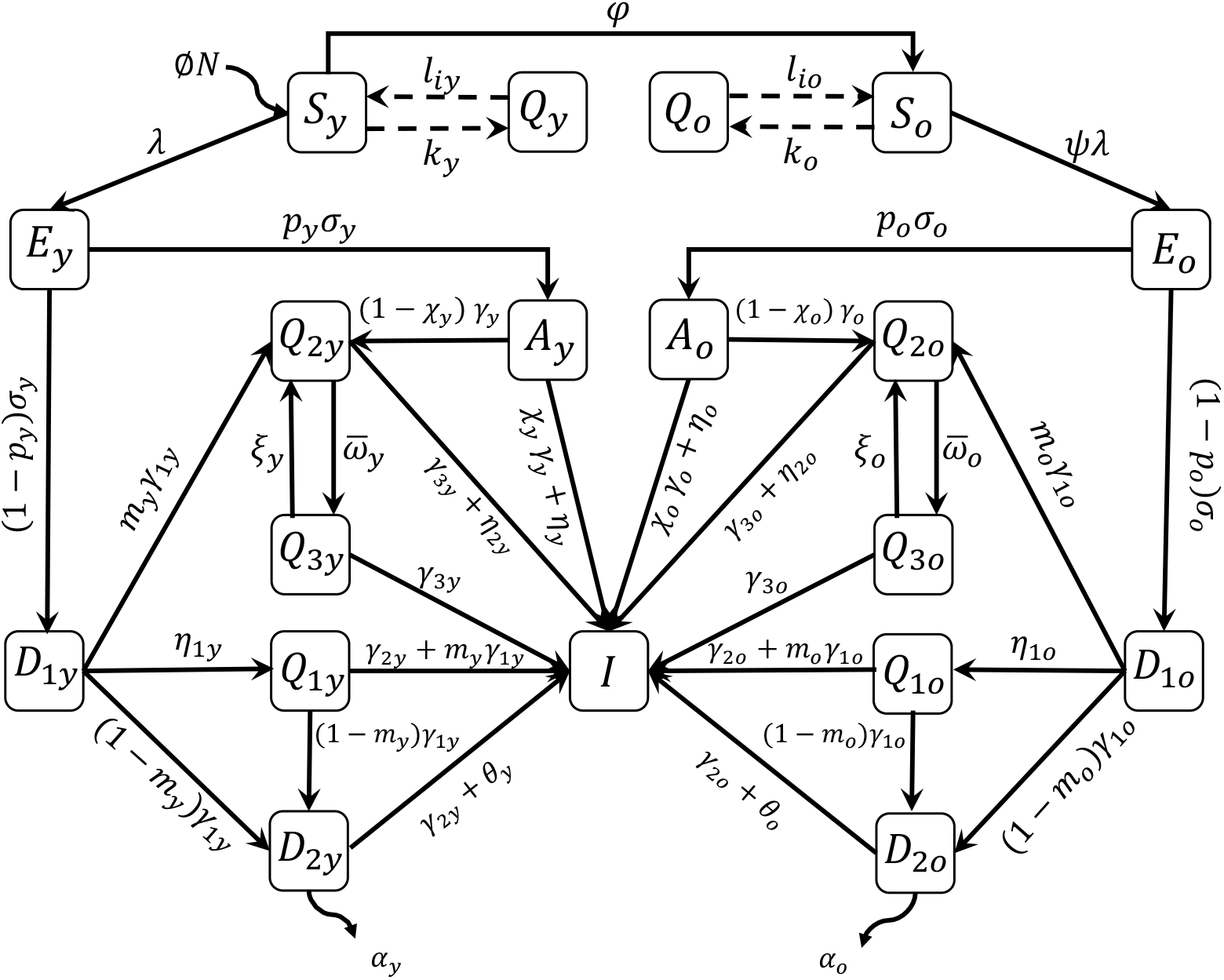
The flowchart of the new coronavirus transmission model.

Equation (2) is simulated permitting intermittent interventions to the boundary conditions. Hence, the new coronavirus transmission model, based on the descriptions of the model variables and parameters summarized in Tables I and II, is described by a system of ordinary differential equations, with *j* = *y*, *o*. Equations for susceptible and isolated persons are

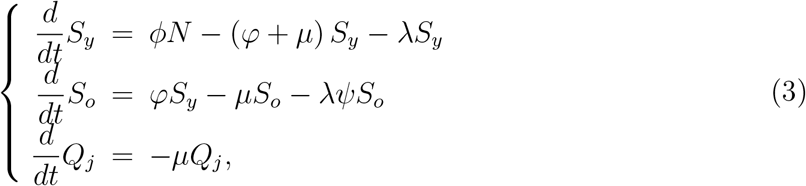

for virus harboring persons,

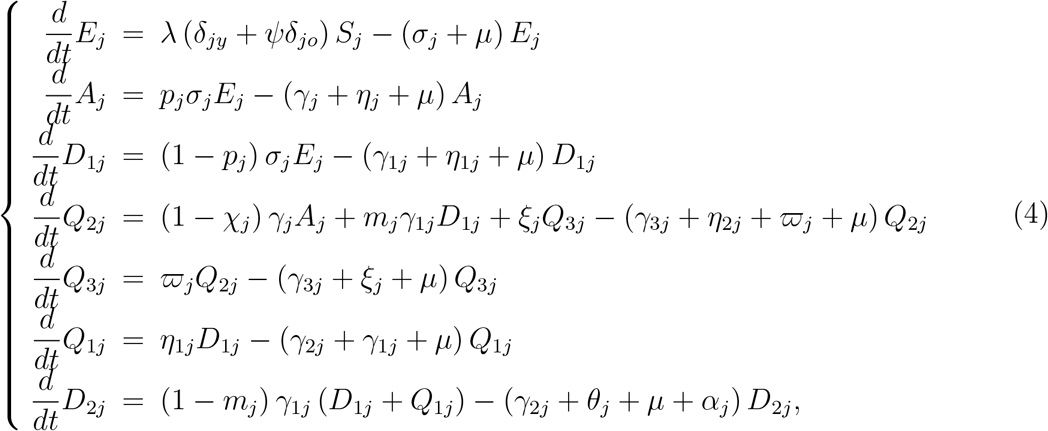

and for immune persons,

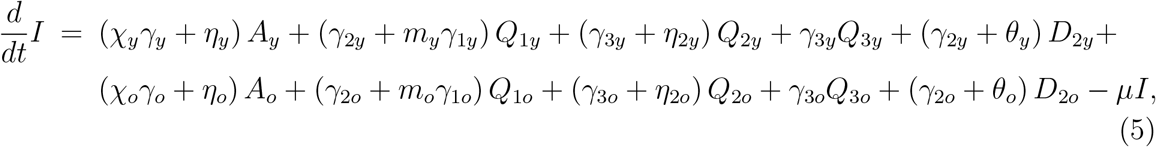

where *N_j_* = *S_j_* + *Q_j_* + *E_j_* + *A_j_* + *D*_1_*_j_* + *Q*_1_*_j_* + *Q*_2_*_j_* + *Q*_3_*_j_* + *D*_2_*_j_*, and *N* = *N_y_* + *N_o_* + *I* obeys

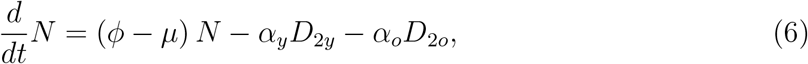

with the initial number of population at *t* = 0 being *N*(0) = *N*_0_ = *N*_0_*_y_* + *N*_0_*_o_*, where *N*_0_*_y_* and *N*_0_*_o_* are the size of young and elder subpopulations at *t* = 0. If *ϕ* = *μ* + (*α_y_D*_2_*_y_* + *α_o_D*_2_*_o_*) /*N*, the total size of the population is constant. The initial and boundary conditions are given in Appendix A.

The number of non-isolated (circulating) susceptible persons *S_j_* is obtained from equation (3), and the number of circulating plus isolated susceptible persons *S^tot^* is obtained by

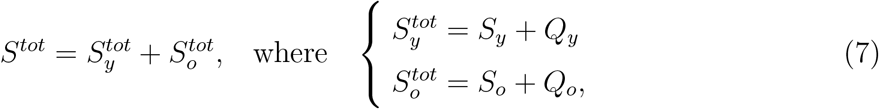

where 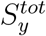 and 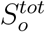 are the numbers of susceptible, respectively, young and elder persons.

From the solution of the system of equations (3), (4), and (5), we can calculate the numbers of accumulated severe covid-19 cases in young (Ω*_y_*) and elder (Ω*_o_*) subpopulations, which are given by the exits from *D*_1_*_y_*, *Q*_1_*_y_*, *D*_1_*_o_*, and *Q*_1_*_o_*, and entering into classes *D*_2_*_y_* and *D*_2_*_o_*, that is,

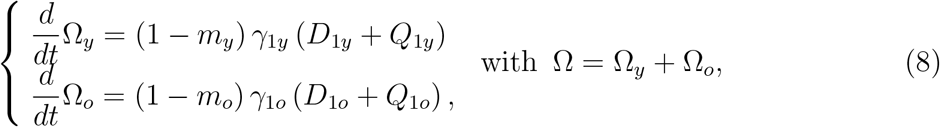

where Ω*_y_*(0) = Ω*_y_*_0_ and Ω*_o_*(0) = Ω*_o_*_0_. The daily severe covid-19 cases Ω*_d_* is, considering Δ*t* = *t_i_* − *t_i_*_−1_ = 1 day,

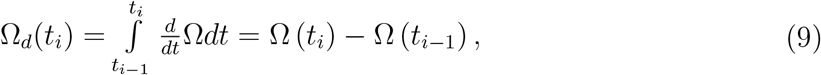

where Ω*_d_*(0) = Ω*_d_*_0_ is the first observed covid-19 case at *t*_0_ = 0, with *i* = 1, 2, ···. The number of deaths due to the severe covid-19 cases is calculated by

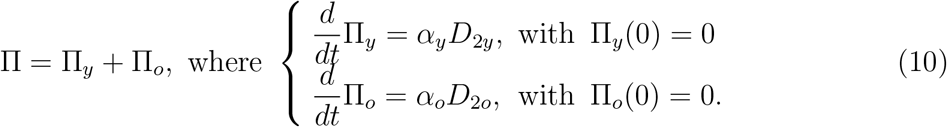

In the estimation of the additional mortality rates, we must bear in mind that the time at which new cases and deaths were registered does not have direct correspondence, rather they are delayed by Δ days, that is, П*_y_* (*t* + Δ) = *α_y_D*_2_*_y_*(*t*), for instance.

### B. The basic and effective reproduction numbers

Among several model variables, we define as the epidemic curve the time-varying number of severe covid-19 cases *D*_2_. This curve, however, is determined by the effective reproduction number *R_ef_*. This number, given by equation (B12) in Appendix B, is written, for the system of equations (3), (4) and (5), as

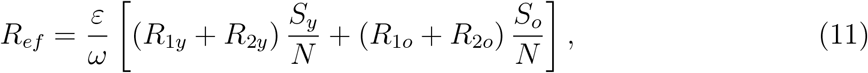

where the partially reduced reproduction numbers *R*_1_*_y_*, *R*_2_*_y_*, *R*_1_*_o_* and *R*_2_*_o_* are given by equation (B8) in Appendix B. The basic reproduction number *R*_0_ is given by equation (B10) in Appendix B letting *ε* = *ω* = 1 (absence of protective measures), *η_j_* = *η*_1_*_j_* = *η*_2_*_j_* = 0 (absence of tests) for *j* = *y*, *o*, and 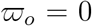 (absence of educational campaign) in *R*_1_*_y_*, *R*_2_*_y_*, *R*_1_*_o_* and *R*_2_*_o_*.

The effective reproduction number *R_ef_* varies during the epidemic as follows.

1. Natural epidemic. The population, initially, did not adopt protection measures against the covid-19 epidemic, hence *ε* = *ω* = 1, and at *t* = 0 we have *R_ef_*(0) = *R*_0_, where the basic reproduction number is obtained by substituting 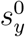 and 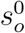 by *N*_0_*_y_*/*N*_0_ and *N*_0_*_o_*/*N*_0_, resulting in

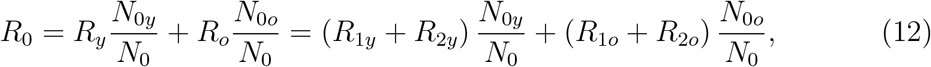

where *R_y_* and *R_o_* are given by equation (B10), and *R*_1_*_y_*, *R*_2_*_y_*, *R*_1_*_o_* and *R*_2_*_o_* are given by equation (B8). For *t* > 0, *R_ef_* decreases as susceptible population decreases.
2. Isolation. At *t* = *τ^is^* a pulse in isolation is introduced, decreasing the number of susceptible persons, from *S_y_* (*τ^is^*^−^) and *S_o_* (*τ^is^*^−^) to *S_y_* (*τ^is^*^+^) and *S_o_* (*τ^is^*^+^), see equations (A3) and (A4) in Appendix A. The decrease in the susceptible populations at *τ^is^* results in *R_ef_*(*τ^is^*^−^) jumping down to *R_ef_* (*τ^is^*^+^) = *R_r_*, where the reduced reproduction number *R_r_* is given by

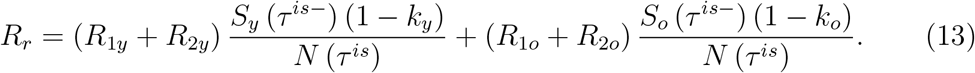 For *t* > *τ^is^*, *R_ef_* decreases as susceptible population decreases.
3. Adoption of protective measures. The protective measures and social distancing are incorporated in the modeling by the factor *ε* in circulating population. At the time of introducing these measures *T*, the effective reproduction number *R_ef_* (*T*_−_) jumps down to *R_ef_* (*T*^+^) = Rp by factor *ε*, that is,

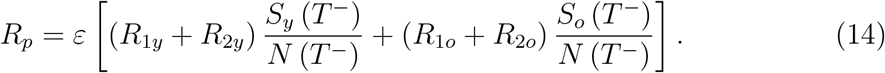 For *t* > *T*, *R_ef_* decreases as susceptible population decreases.
4. Interiorization of the epidemic. The transmission rates in small cities are lower than in larger cities, which is described by the reduction factor *ω*. At the time of the interiorization of epidemic *T_in_*, the effective reproduction number 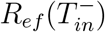 jumps down to 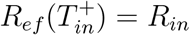 by factor 1/*ω*, that is,

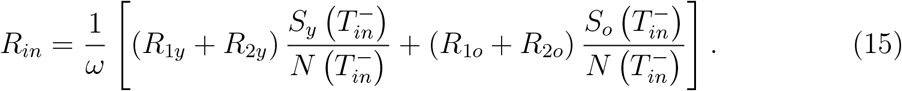 For *t* > *T_in_*, *R_ef_* decreases as susceptible population decreases.
5. Release. An *i*-th release in pulse occurs at time *t_i_* increasing the number of susceptible persons, from 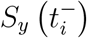 and 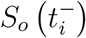 to 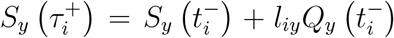 and 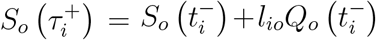. The increase in the susceptible populations at *t_i_* results in 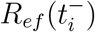 jumping up to 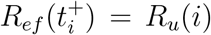, where the increased reproduction number 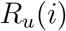 is given by

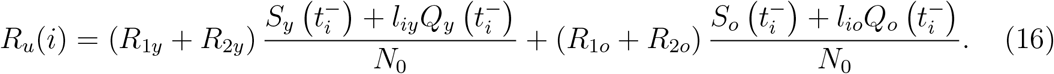 For *t* > *t_m_*, *R_ef_* decreases as susceptible population decreases, and when *t* → *∞*, we have *R_ef_* = 1, and *s*^*^ = 1/*R*_0_.

Notice that at the time of the release, the protective measures can be changed. In this case, the sum of jumping up *R_u_*(*i*) at the *i*-th release and jumping down *R_p_* due to protective measures can result in the increase or decrease in the overall *R_ef_*.

The epidemic curve associated with the steps 1 to 4 describes the isolation, while step 5 is associated with the epidemic under the release. Hence, the knowledge of *R_ef_* during the epidemic can help to plan strategies for the release.

### C. Evaluating the clinical evolution, deaths, and cures

There are three stages in the clinical evolution of covid-19 [2] and [7]. From equation (8), we can derive the number of persons in viraemia *B*_1_ (phase 1), the number of patients with inflammatory response needing hospital care *B*_2_ (phase 2), and the number of patient with cytokine storm needing ICUs/intubated care *B*_3_ (phase 3). From the numbers of *B*_1_, *B*_2_, and *B*_3_, we obtain the number of deaths due to covid-19, and the number of cured persons.

The number of persons in phase 1 is *B*_1_ = *B*_1_*_y_* + *B*_1_*_o_*, for *j* = *y*, *o*, where

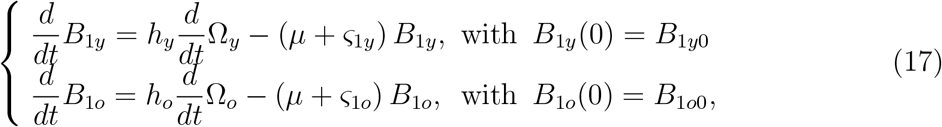

with *d*Ω*_j_*/dt being given by equation (8). The number of persons needing hospital care (inpatients) in phase 2 is *B*_2_ = *B*_2_*_y_* + *B*_2_*_o_*, where

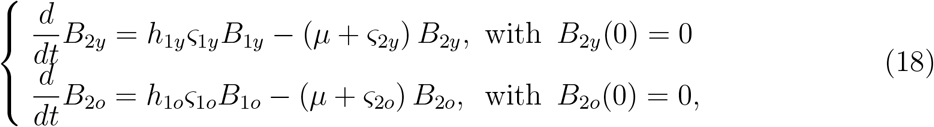

and the number of persons needing ICUs/intubated care (ICUs patients) in phase 3 is *B*_3_ = *B*_3_*_y_* + *B*_3_*_o_*, where

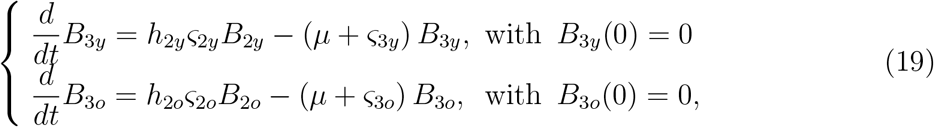

with the total number of severe covid-19 cases being *B* = *B*_1_ + *B*_2_ + *B*_3_.

The number of deaths caused by severe covid-19 cases can be calculated from the fatality associated with the clinical evolution divided into *B*_1_, *B*_2_, and *B*_3_. The number of deaths of persons in phase 1 is П_1_ = П_1_*_y_* + П_1_*_o_*, where

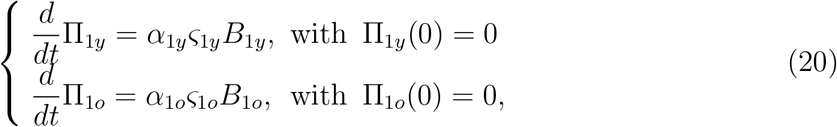

the number of deaths of inpatients in phase 2 is П_2_ = П_2_*_y_* + П_2_*_o_*, where

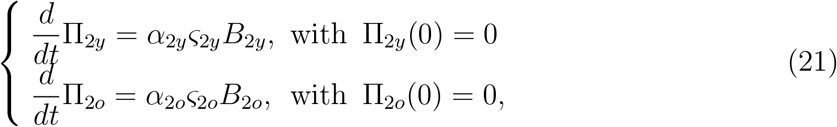

and the number of deaths of ICUs patients in phase 3 is П_3_ = П_3_*_y_* + П_3_*_o_*, where

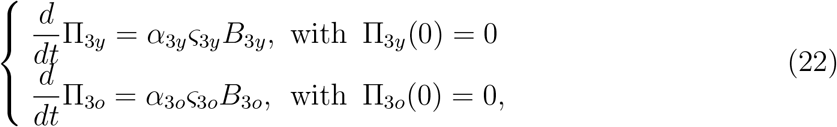

with the total number of deaths being П*_s_* = П_1_ + П_2_ + П_3_.

The total number of persons being cured is *C* = *C_y_* + *C_o_*, where

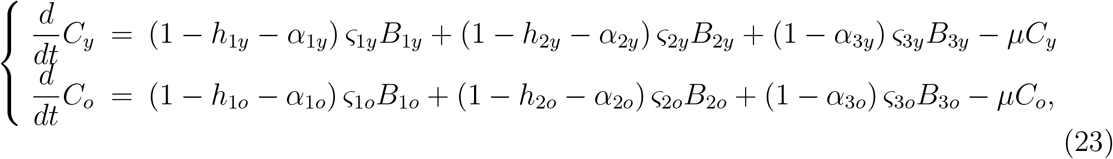

with *C_y_* (0) = *C_y_* (0) = 0.

Concerning the parameters of *B*, П, and *C*, for *j* = *y*, *o*, a proportion *h_j_* of pre-disease persons enter into phase 1 (non-hospitalized, class *B*_1_), from which a proportion *h*_1_*_j_* enters to phase 2 (inpatients, class *B*_2_), and a proportion *h*_2_*_j_* enters to phase 3 (ICUs patients, class *B*_3_). The average spending time of persons in phases 1, 2, and 3 are, respectively, 1/ *ς*_1_*_j_*, 1/ *ς*_2_*_j_*, and 1/ *ς*_3_*_j_*, where *ς*_1_*_j_* and *ς*_2_*_j_* are the disease progression rates, respectively, from phase 1 to 2, and phase 2 to 3, and *ς*_3_*_j_* is the rate of exiting from phase 3. The additional mortality (fatality) proportions among patients in phases 1,2, and 3 are, respectively, *α*_1_*_j_*, *α* _2_*_j_*, and *α* _3_*_j_*. The fraction 1 − *h_j_* is the non-hospitalization of pre-disease covid-19 cases, and 1 − h_1_*_j_* − *α*_1_*_j_*, 1 − *h*_2_*_j_* − *α*_2_*_j_*, and 1 − *α*_3_*_j_* are the proportions of cure of patients in phases 1, 2, and 3.

Table III summarizes the parameters related to the clinical evolution and values (for elder classes, values are between parentheses), see Appendix C.

**TABLE III:**
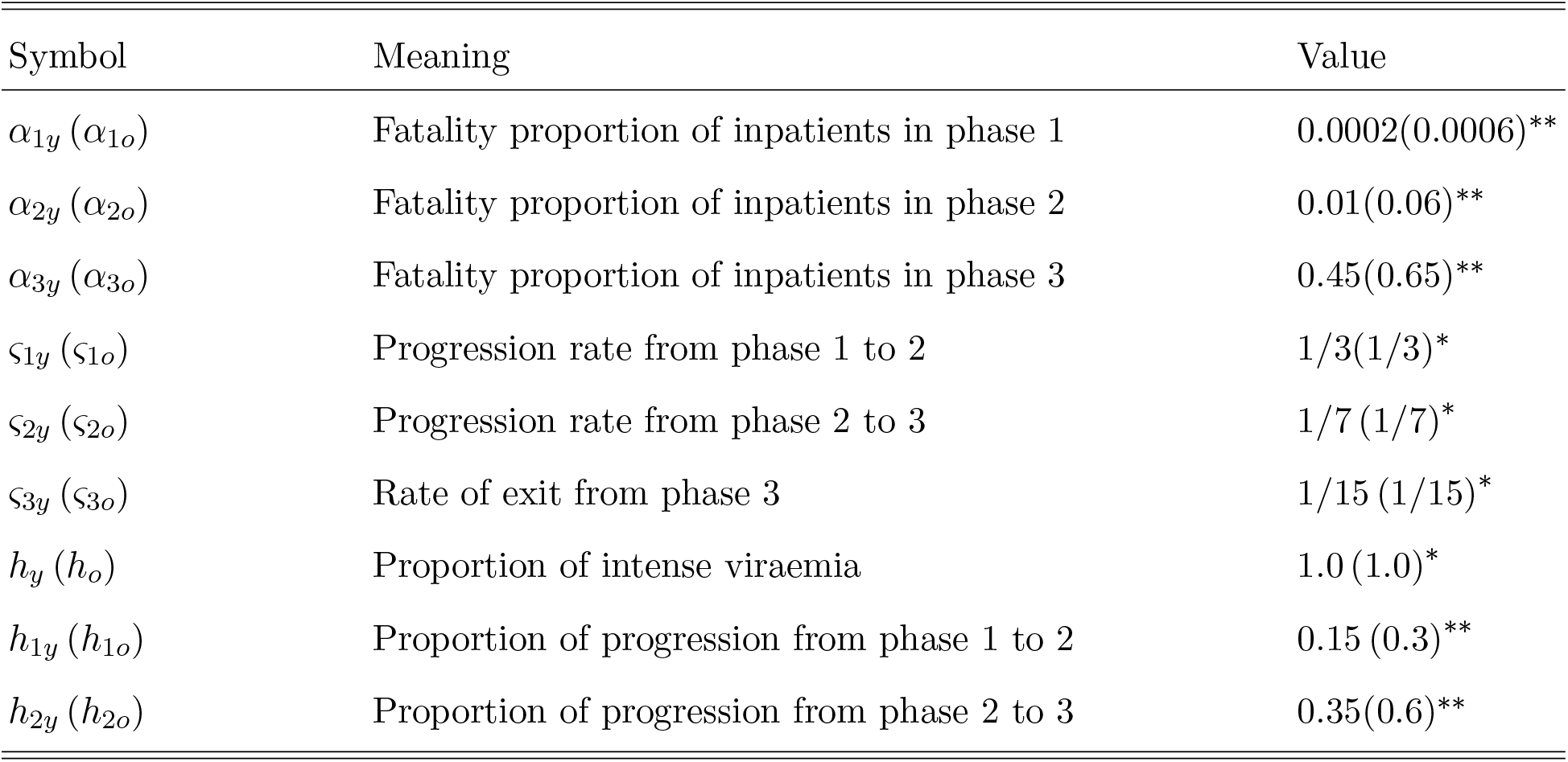
Summary of the parameters related to hospitalization (*j* = *y*, *o*) and values (rates in days^−1^ and proportions are dimensionless). Some values are assumed (^*^) or estimated (^**^).

## III. RESULTS

On March 24, São Paulo State implemented the isolation of the persons in non-essential activities until June 15. Based on the analysis of the model presented in the foregoing section, we evaluate the scenarios of isolation and further release.

### A. Epidemiological scenario of isolation

In [4], we estimated the model parameters to describe the epidemiological status of the new coronavirus in the population in isolation in São Paulo State. Additionally, the protective measures adopted by the population reduced the transmission of infection beyond that reduced by isolation. However, the decreased populational density and contact rate in the cities out of the metropolitan region of São Paulo City contributed to diminishing, even more, the epidemic of covid-19 in São Paulo State. These small and medium cities having 52.6% of inhabitants [8] contributed to reducing the overall epidemic (we called the interiorization of the epidemic), which is estimated here. We assumed that the epidemic was not occurring in the isolated population, hence the scenario is the epidemic with interventions in the circulating population.

Let us summarize the estimated values of the model parameters using data from February 26 to July 1 [8], where the subscripts *y* and *o* stand for, respectively, young and elder subpopulations (see Appendix C).

1. The transmission rates, from February 26 to April 3– Estimated values were *β_y_* = 0:78 and *β_o_* = 0:90 (both in *days*^−1^), where *ψ* = 1:15, resulting in the basic reproduction number *R*_0_ = 9:24 (partials *R*_0_*_y_* = 7:73 and *R*_0_*_o_* = 1:51).
2. The proportion in isolation, from March 24 to April 12 { Estimated value was *k* = *k*_mean_ = 0:53. The effects of the isolation appear 9 days later (see [4]).
3. The protective factor, from April 4 to May 7 – Estimated value was *ε* = 0:5. The transmission rates *β_y_* = 0:78 and *β_o_* = 0:90 (both in *days*^−1^) are reduced to 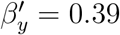 and 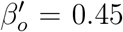 (both in *days*^−1^), resulting in the reduced basic reproduction number 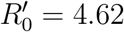 (partials 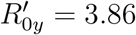 and 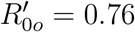).
4. The additional mortality rates, from March 16 to May 13 – Fixing Δ = 15 *days* and Γ = 0.26 (74% of deaths are occurring in elder persons with severe covid-19), estimated values were *α_y_* = 0.00185 and *α_o_* = 0.0071 (both in *days*^−1^). The death occurs 15 days after the onset of severe covid-19 symptoms (see [4]).
5. The reduction factor by interiorization of the epidemic, from May 1 to July 1 – Estimated value is ω = 1.1. The transmission rates 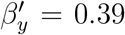 and 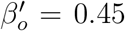 (both in *days*^−1^) are reduced to 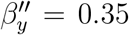 and 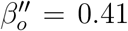 (both in *days*^−1^), resulting in the reduced basic reproduction number 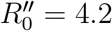 (partials 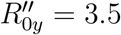 and 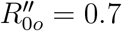).

Details of the estimation in steps 1 to 4 can be found in [4], and the estimation of step 5 is done here using equation (C1) in Appendix C. Figures 1-2 summarize the above estimations.

**FIG. 1:**
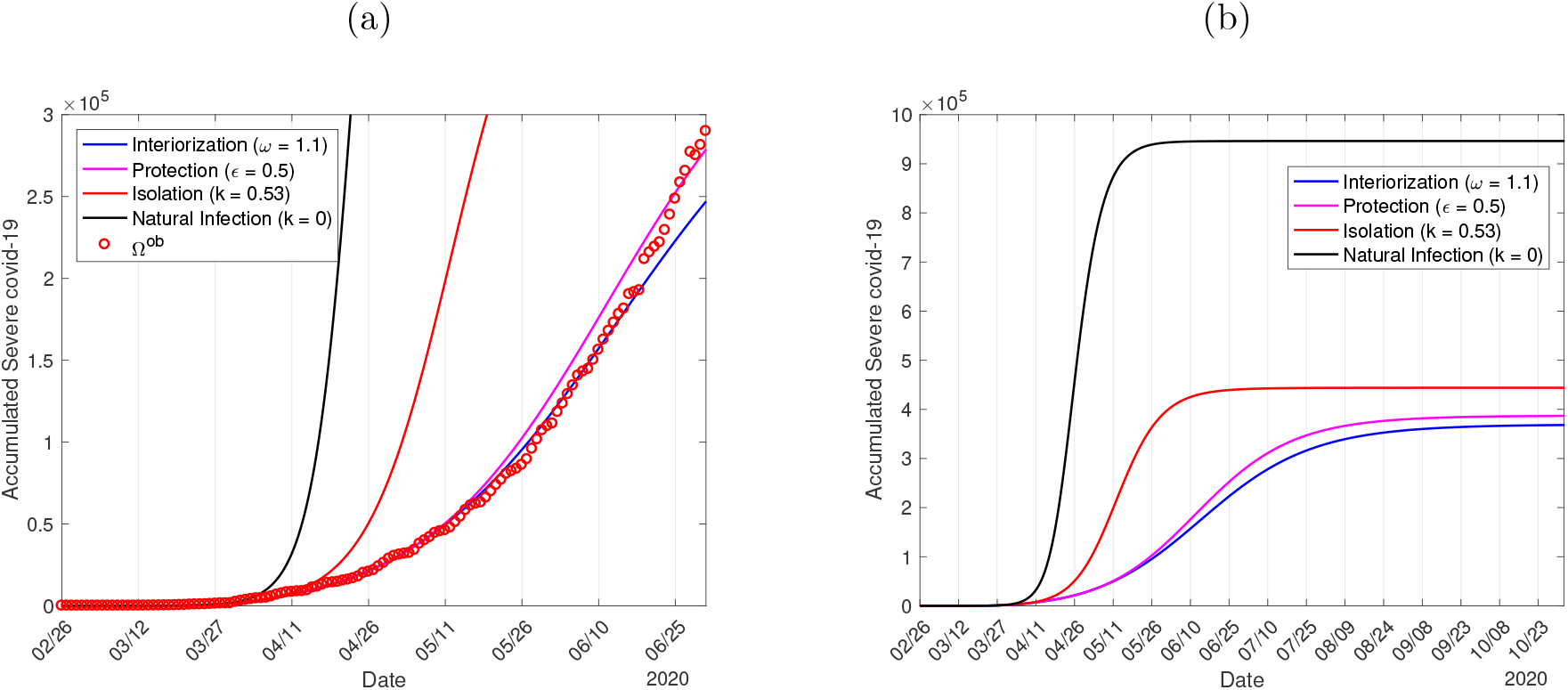
The curves of natural epidemic (*k* = 0), epidemic with isolation (*k* = 0.53), epidemic occurring with isolation and protective measures (*k* = 0.53 and *ε* = 0.5), and epidemic with interiorization (*k* = 0.53, *ε* = 0.5, and *ω* = 1.1) (a), and the extended curves

**FIG. 2:**
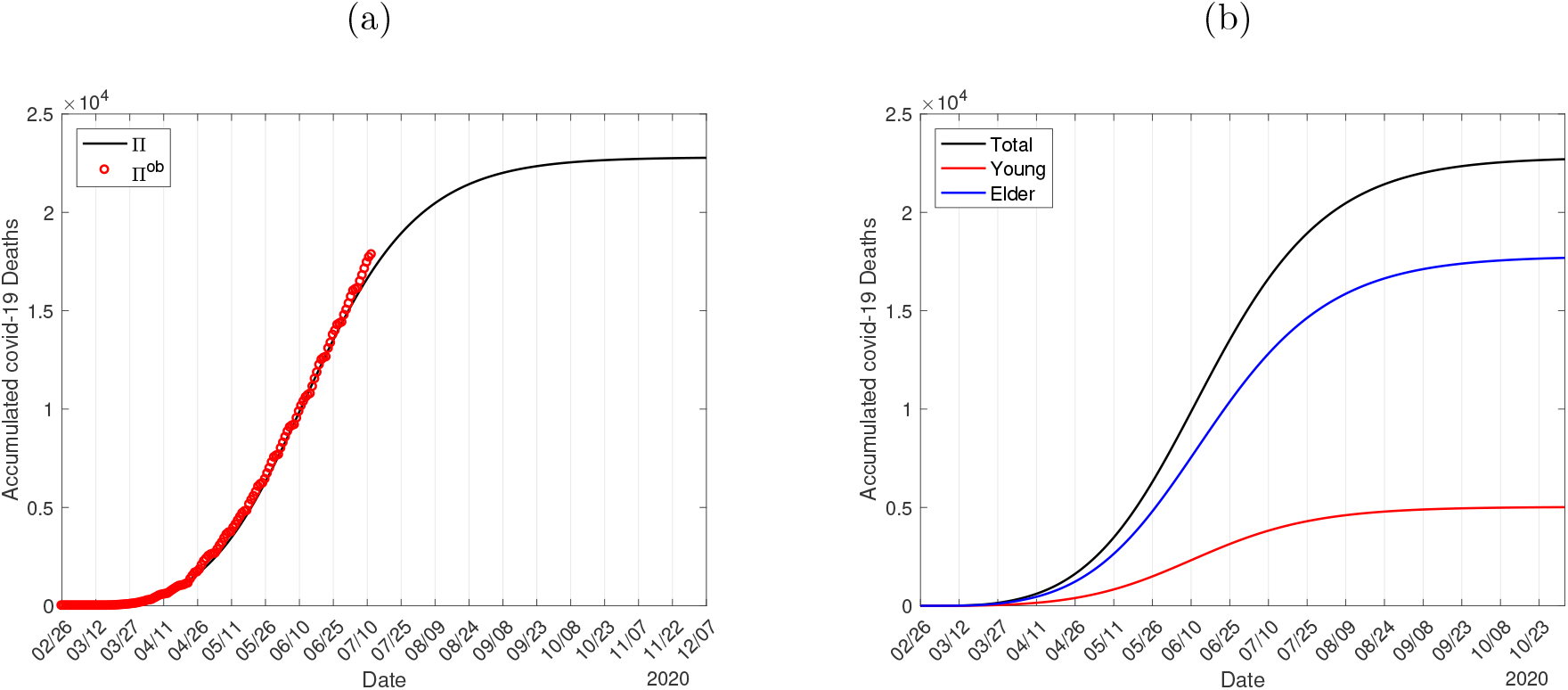
The estimated curves of the accumulated deaths due to covid-19 П with Δ = 15 *days*, and the observed data (a), and the extended curves for young П*_y_*, elder П*_o_*, and total П = П*_y_* + П*_o_* persons (b).

Figure 1 shows the estimated curves of Ω and the observed data (a), and the extended curves for young (Ω*_y_*), elder (Ω_0_), and total (Ω = Ω*_y_* + Ω_0_) persons (b). The curve of Ω is given by equation (8).

At the end of the first wave of the epidemic, for the natural epidemic, the numbers of accumulated severe covid-19 cases for Ω*_y_*, Ω*_o_*, and Ω = Ω*_y_* + Ω_0_ are, respectively, 605, 300, 341,100, and 946, 400, and the numbers of immune young (*I_y_*), elder (*I_o_*), and total (*I* = *I_y_* + *I_o_*) persons are, respectively, 37.5 million, 6.7 million, and 44.2 million. For the epidemic with interventions, from Figure 1(b), the numbers of accumulated severe covid-19 cases for Ω*_y_*, Ω*_o_*, and Ω = Ω*_y_* + Ω_0_ are, respectively, 231, 400 (38%), 136, 700 (40%), and 368,100 (39%), while the numbers of *I*_y_, I_o_, and I = *I_y_* + I_o_ are 14.4 million (38.4%), 2.7 million (40.3%), and 17.1 million (38.7%). The percentage between parentheses is the ratio with respect to the natural epidemic.

In the natural epidemic, the percentage of severe covid-19 cases for young and elder subpopulations are, respectively, 64% and 36%, with young subpopulation having 1.8-time more cases than elder subpopulation, while in epidemic with interventions, we have 63% and 37%, with young subpopulation having 1.7-time more cases than elder subpopulation. However, São Paulo State has 15.3% of the elder population, that is, young subpopulation has 5.5-time more inhabitants than elder subpopulation. This result suggests that elder subpopulation has 3-time more risk of infection when in interaction with young subpopulation.

On June 16 we observe the first point detached from the curve, showing that on June 7, 9 days earlier, some factors changed the epidemic curve. If the proportion in isolation decreased, then the number of severe covid-19 cases must increase since June 16, and the number of deaths must also increase after 15 days, that is, since June 22. If the number of deaths does not increase since June 22, then the increase can be explained by the increased number of tests in mild covid-19 cases.

Figure 2 shows the estimated curves of П and the observed data (a), and the extended curves of deaths for young (П*_y_*), elder (П*_o_*), and total (П = П*_y_* + П*_o_*) persons (b). The curve of n is given by equation (10). We used Δ = 15 *days*, which comes from the value of 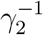 being 12 for young and 21 for elder persons, remembering that elder persons are under a higher fatality rate than young persons. On June 30, we observe the first point quite detached from the curve, showing that on June 15, 15 days earlier, some factors changed the epidemic curve.

Equation (C2) in Appendix C evaluates the number of deaths from severe covid-19 cases *D*_2_. The average period in class *D*_2_ is (*γ*_2_*_j_* + *θ_j_* + *μ* + *α_j_*)^−1^, for *j* = *y*, *o*, and the probability of death is *α_j_*/ (*γ*_2_*_j_* + *θ_j_* + *μ* + *α_j_*). Using the values given in Table II, the probabilities of fatality induced by severe covid-19 for young and elder persons are, respectively, 2% and 13%. At the end of the first wave of the natural epidemic, the accumulated numbers of deaths for П*_y_*, П*_o_*, and П = П*_y_* + П*_o_* are, respectively, 13,110, 44, 230, and 57, 340. Notice that these numbers of deaths were calculated using the estimated fatality rates *α_y_* = 0.00185 and *α_o_* = 0.0071 (both in *days*^−1^), that is, we did not account for the deaths of severe covid-19 cases occurring in patients without hospital care. For the epidemic with interventions, from Figure 2(b), the accumulated numbers of deaths for П*_y_*, П*_o_*, and П = П*_y_* + П*_o_* are, respectively, 5,000 (38%), 17, 700 (40%), and 22, 700 (39.5%). The percentage between parentheses is the ratio with respect to the natural epidemic.

From Figures 1-2, at the end of the first wave of the epidemic, the isolation of 53% in the population, the reduction in 50% by protective measures and decrease in transmission rates by *ω* = 1.1 resulted in 60% of reduction in the numbers of severe covid-19 cases, immunes and deaths. Moreover, at the end of the first wave of the epidemic, the susceptible persons in the circulating population are *S_y_* = 3.7 million, *S_o_* = 0.5 million, and *S* = *S_y_* + *S_o_* = 4.2, which summed with the isolated population result in 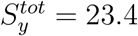 million, 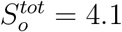 million, and 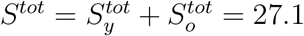 million, from equation (7). Hence, at the end of the first wave of the epidemic, the remaining susceptible and immune persons are, respectively, 61% and 38% of the entire population of São Paulo State (44.6 million), showing that the risk of rebound of the epidemic with the release of isolated persons is higher.

Figure 3 shows the calculated curve of Ω*_d_* and the observed daily cases (a), and the initial part of the estimated curve of Ω with observed data Ω*^ob^*, the extended Ω*_d_* and daily observed cases 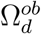, and severe cases *D*_2_ (b). The curve of 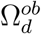 is given by equation (9), and *D*_2_ is solution of the system of equations (3), (4) and (5). The peaks of the daily Ω*_d_* and severe *D*_2_ cases are, respectively, 4, 468 and 57,900, occurring on June 14 and June 26. The peaks of *D*_2_ for young and elder persons are, respectively, 30,800 and 27,100, occurring on June 25 and 27.

**FIG. 3:**
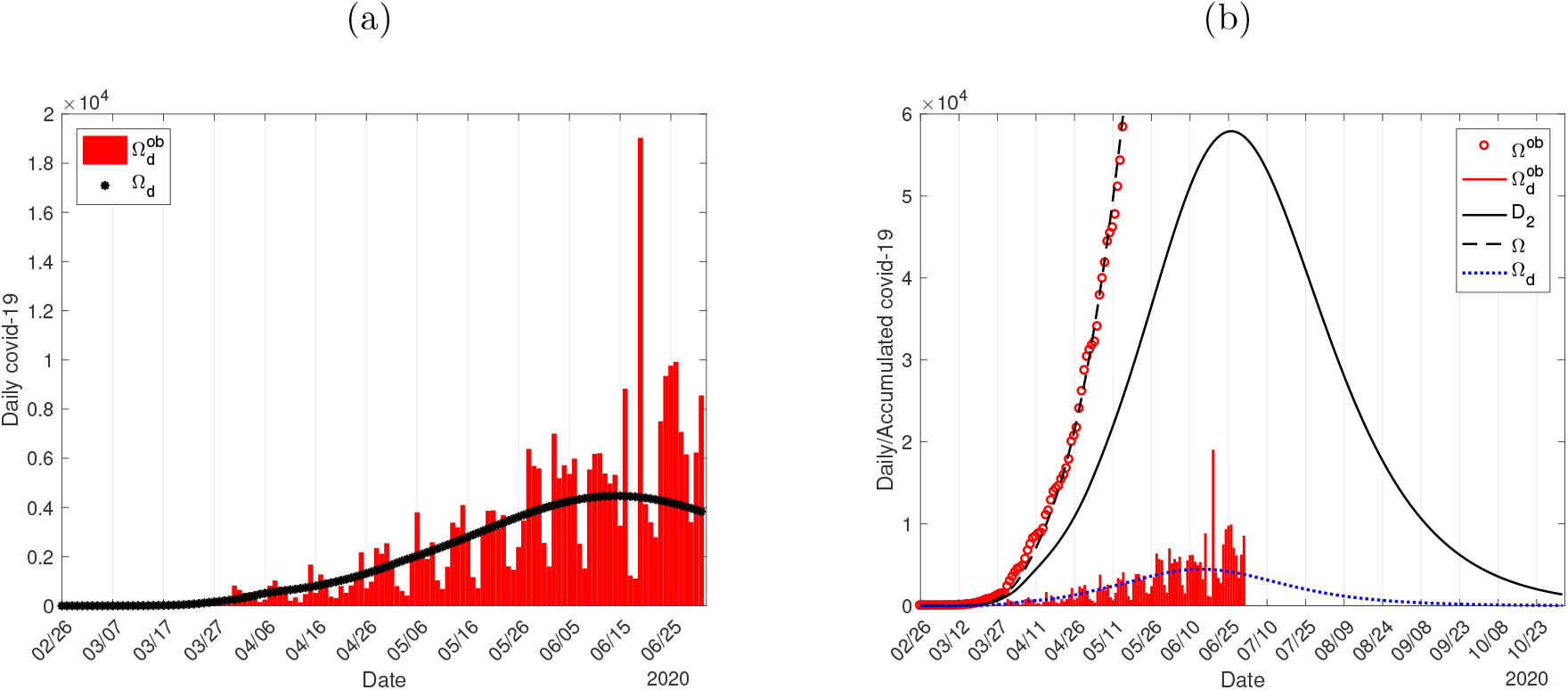
The calculated curve of Ω*_d_* and the observed daily cases (a), and the initial part of the estimated curve of Ω with observed data Ω*^ob^*, the extended curve of Ω*_d_* and daily observed cases 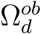, and severe covid-19 cases *D*_2_ (b).

Figure 4 illustrates the effective reproduction number *R_ef_* and D_2_ during the natural epidemic (a), and the epidemic with interventions (b). To be fitted together in the same frame with *R_ef_*, the curve of *D*_2_ is divided by 40, 000 in Figure 4(a) and 6,000 in Figure 4(b). At the peak of the epidemic, the effective reproduction number is lower than one, hence we have *R_ef_* = 1 occurring on April 6 for the natural epidemic, and on June 16 for the epidemic with interventions.

**FIG. 4:**
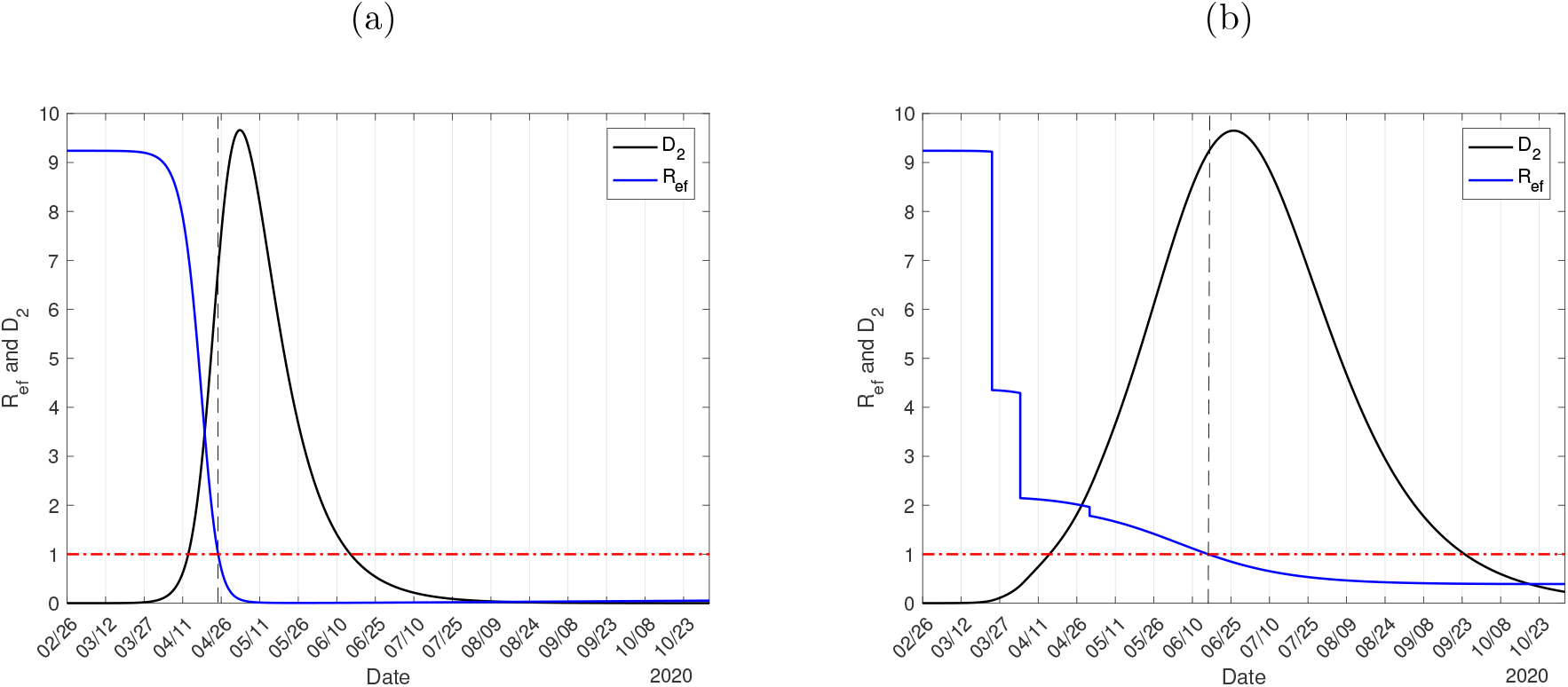
The effective reproduction number *R_ef_* for natural epidemic (a), and epidemic with interventions and interiorization (b). The number of severe covid-19 cases *D*_2_ must be multiplied by 40, 000 (a) and 6, 000 (b).

From Figure 4(a), the effective reproduction number *R_ef_* decreases quickly and approaches to value near zero at the beginning of July. Hence, the natural epidemic curve increases and decreases quickly, and the majority of severe covid-19 cases occur during approximately two months when *R_ef_* > 1, which definitively results in the collapse of the health care system. From Figure 4(b), at the beginning of the epidemic, on February 26, we have *R_ef_* = *R*_0_ = 9.24, on March 24, a jump down occurred to *R_ef_* = 4.35 due to the isolation, and a new jump down occurs to *R_ef_* = 2.15 on April 4 when protective measures were adopted. On May 1 another small jump down to *R_ef_* = 1.78 occurs due to interiorization. On June 15, when the release began, we have *R_ef_* = 1.02, but in the ascending phase of the epidemic. At the end of the first wave of the epidemic, we have *R_ef_* = 0.39 achieved already in September, but with elevate number of infected persons. The knowledge of *R_ef_* could help public health authorities to plan the strategies of release.

Figure 3(b) showed the number of severe covid-19 cases *D*_2_ calculated from the estimated accumulated cases Q shown in Figure 1. From *D*_2_, the number of accumulated deaths shown in Figure 2 was obtained. Instead of non-specified severe covid-19 cases *D*_2_, we use the clinical evolution since the onset of symptoms. Hence, we evaluate the number of patients in phases 1 (*B*_1_), 2 (*B*_2_), and 3 (*B*_3_), and the number of deaths is obtained taking into account these three phases of covid-19.

Figure 5 shows the calculated curves of *B*_1_, *B*_2_, *B*_3_, and *B* = *B*_1_ + *B*_2_ + *B*_3_, for young (a) and elder (b) subpopulations. We used the values given in Table III, and equations (17), (18) and (19) for *B*_1_, *B*_2_, and *B*_3_. For young persons, the peaks of *B*_1_, *B*_2_, and *B*_3_ are 8, 328, 2, 843, and 1, 957, occurring on June 17, 24, and July 7, while for elder persons, the peaks are 5, 015, 3, 422, and 4, 030, occurring on June 15, 22, and July 5. The peaks of *B* for young, elder, and total persons are, respectively, 12, 840, 12, 050, and 24, 890, occurring on June 21, 23, and 22.

**FIG. 5:**
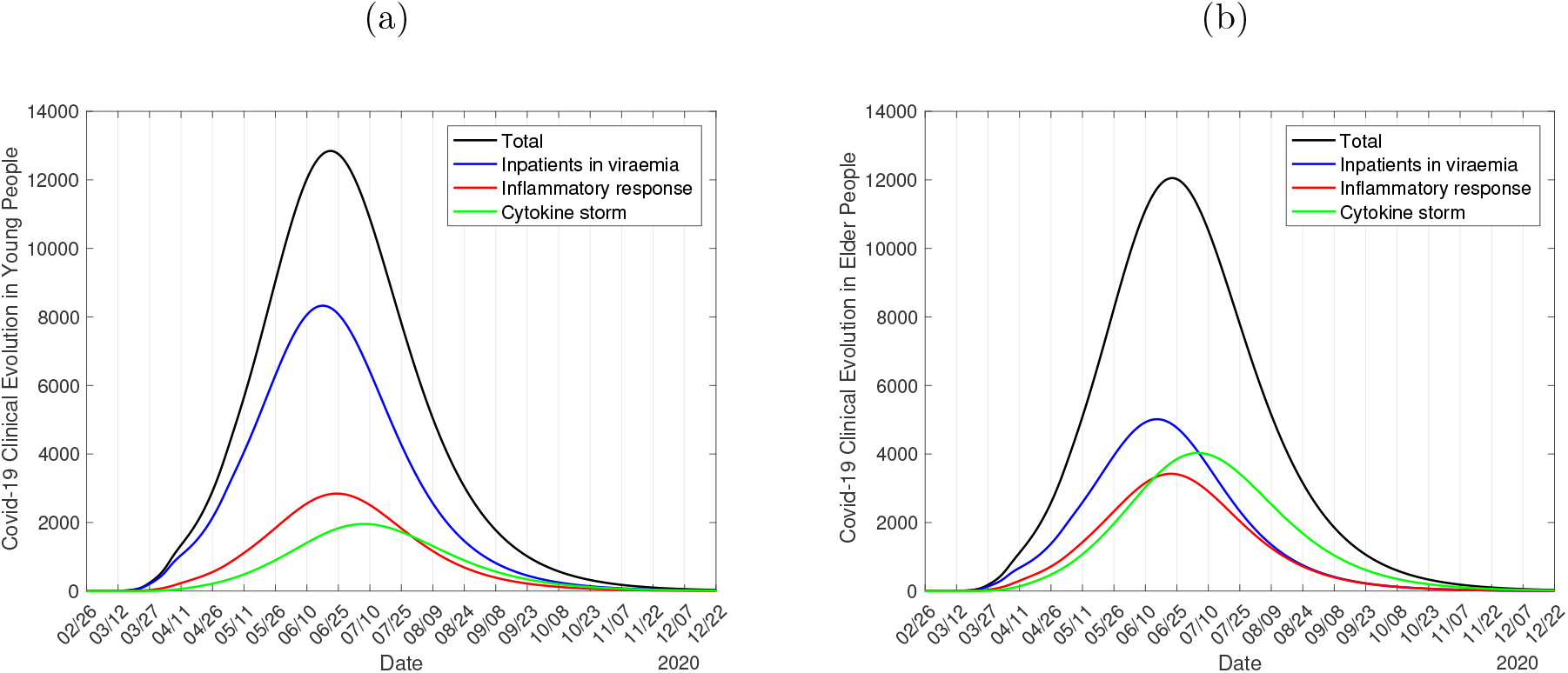
The calculated curves of *B*_1_, *B*_2_, *B*_3_ and *B* = *B*_1_ + *B*_2_ + *B*_3_, for young (a) and elder (b) subpopulations.

From Figure 3, the peaks of daily Ω*_d_* and severe *D*_2_ cases occurred, respectively, on June 14 and 26. From Figure 5, we retrieve the values of *B*_2_ = *B*_2_*_y_* + *B*_2_*_o_* (inpatients) and *B*_3_ = *B*_3_*_y_* + *B*_3_*_o_* (ICUs patients) for some dates to compare with those registered in São Paulo State. On June 14, the numbers of inpatients and ICUs patients are 5, 990 and 4, 834, totalizing 10, 824. On June 26, the numbers of inpatients and ICUs patients are 6, 235 and 5, 735, totalizing 11, 970. On July 10, the numbers of inpatients and ICUs patients are 5, 413 and 5, 937, totalizing 11, 350. On July 14, the numbers of inpatients and ICUs patients are 4, 988 and 5, 814, totalizing 10, 802. Summarizing, on June 26 we have the higher number of inpatients (6, 235), and on July 10, the higher number of ICUs patients (5, 937).

In Sao Paulo State [9], on June 14, the numbers of inpatients (*B*_2_) and ICUs patients (*B*_3_) are 8, 272 and 5, 710, totalizing 13, 982. On June 26, the numbers of *B*_2_ and *B*_3_ are 8, 274 inpatients and 5,666 ICUs patients, totalizing 13, 940. On July 10, the numbers of *B*_2_ and *B*_3_ are 8, 259 inpatients and 5, 291 ICUs patients, totalizing 13, 550. On July 14, the number of *B*_2_ and *B*_3_ are 9,116 inpatients and 6,173 ICUs patients, totalizing 15, 289. Notice that the numbers of inpatients and ICUs patients maintained practically constant, except for an increase on July 14. The number of inpatients is higher than that predicted by the model, while the number of ICUs patients is close. A possible explanation is the absence of tests among inpatients at the moment of entering to receive hospital care, while the patients needing ICUs patients may have been tested positive for covid-19.

In the natural epidemic, the peak of severe covid-19 cases is 386, 400 occurring on May 3, while for the epidemic with interventions, the peak is 57, 900 occurring on June 26. From Figure 5, on June 26, we have 6, 235 inpatients and 5, 735 ICUs patients, totalizing 11, 970 (21% of the peak of severe cases). In a naive comparison assuming proportionality, for the natural epidemic on May 3, we could have 40, 254 inpatients and 37, 026 ICUs care, totalizing 77, 280. These values definitively show the collapse of the health care system, and the surplus number of patients who did not receive treatment could have died, and the total number of deaths increases beyond that caused by the fatality of covid-19.

Figure 6 shows the estimated curve of isП_s_ with 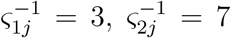 and 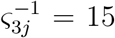, *j* = *y*, *o* (all in *days*), summing 25 *days*) and the observed daily cases of death (a), and the curves of *C_y_* and *C_o_* for young (continuous curve) and elder (dashed curve) subpopulations (b). In Figure 6(a) we show also two curves using Δ_1_ = 15 and 25 *days*. We used equation (C3) in Appendix C to estimate the parameters given in Table III, which were used to obtain the curves of *C_y_* and *C_o_* by equation (23). At the end of the first wave of the epidemic, the accumulated cure for young and elder persons are, respectively, 224, 600 and 117, 600.

**FIG. 6:**
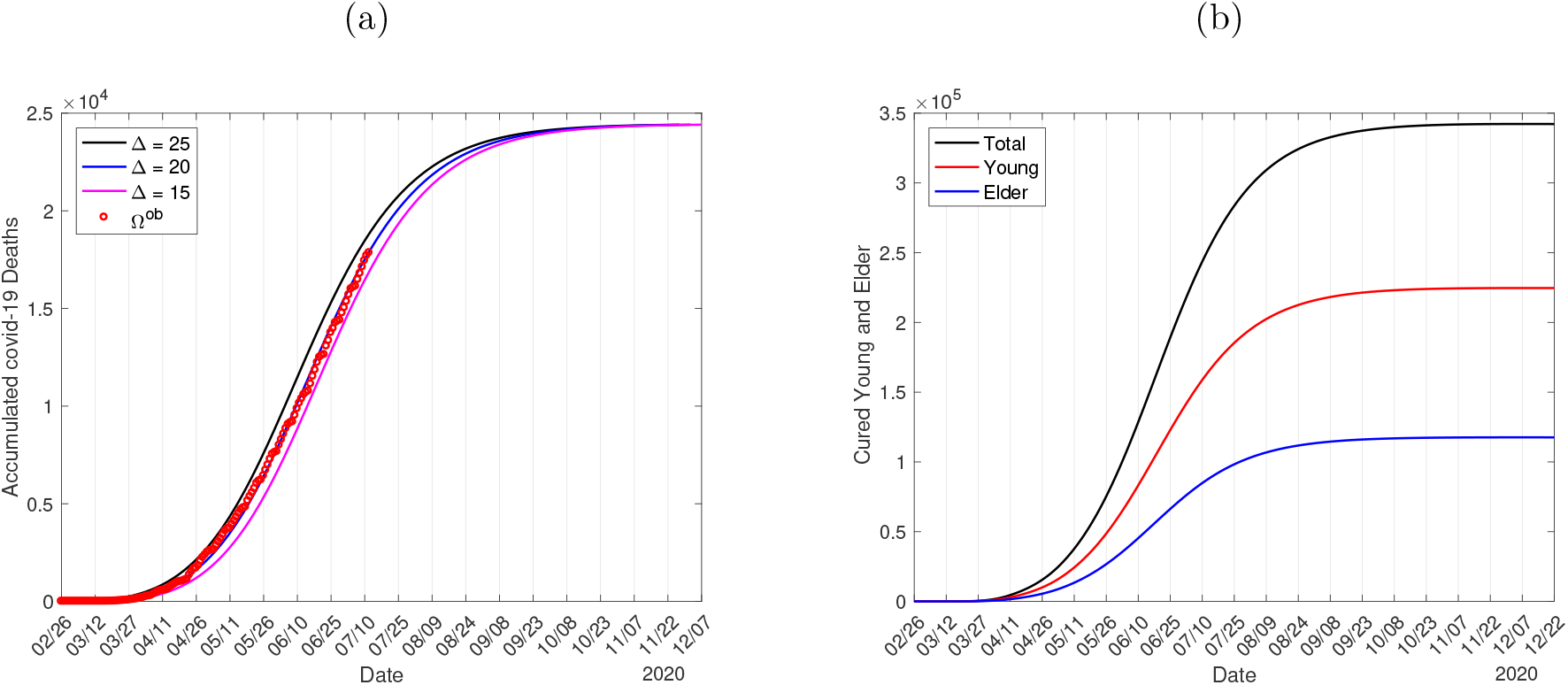
The estimated curve of П*_s_* with Δ = 20 *days*, plus curves for Δ = 15 and 25 *days*, and the observed daily cases of death (a), and the curves of cured persons *C_y_*, *C_o_* and *C* = *C_y_* + *C_o_*(b).

Based on Figure 6(a), Figure 7 shows the curves of П_1_, П _2_, П _3_, and П*_s_* = П_1_ + П_2_ + П_3_ for young (a) and elder (b) subpopulations. We used the values given in Table III, and equations (20), (21) and (22) for П_1_, П_2_, and П_3_. At the end of the first wave of the epidemic, for young persons, the values of П_1_, П_2_, and П_3_, are 46, 348, and 5, 471, while for elder persons, the values are 82, 2, 465, and 16, 000. The accumulated deaths for young, elder, and total persons are, respectively, 5,865 (2.5%), 18, 547 (13.6%), and 24, 412 (6.6%). The percentage between parentheses is the severe covid-19 case fatality rate П/Ω.

**FIG. 7:**
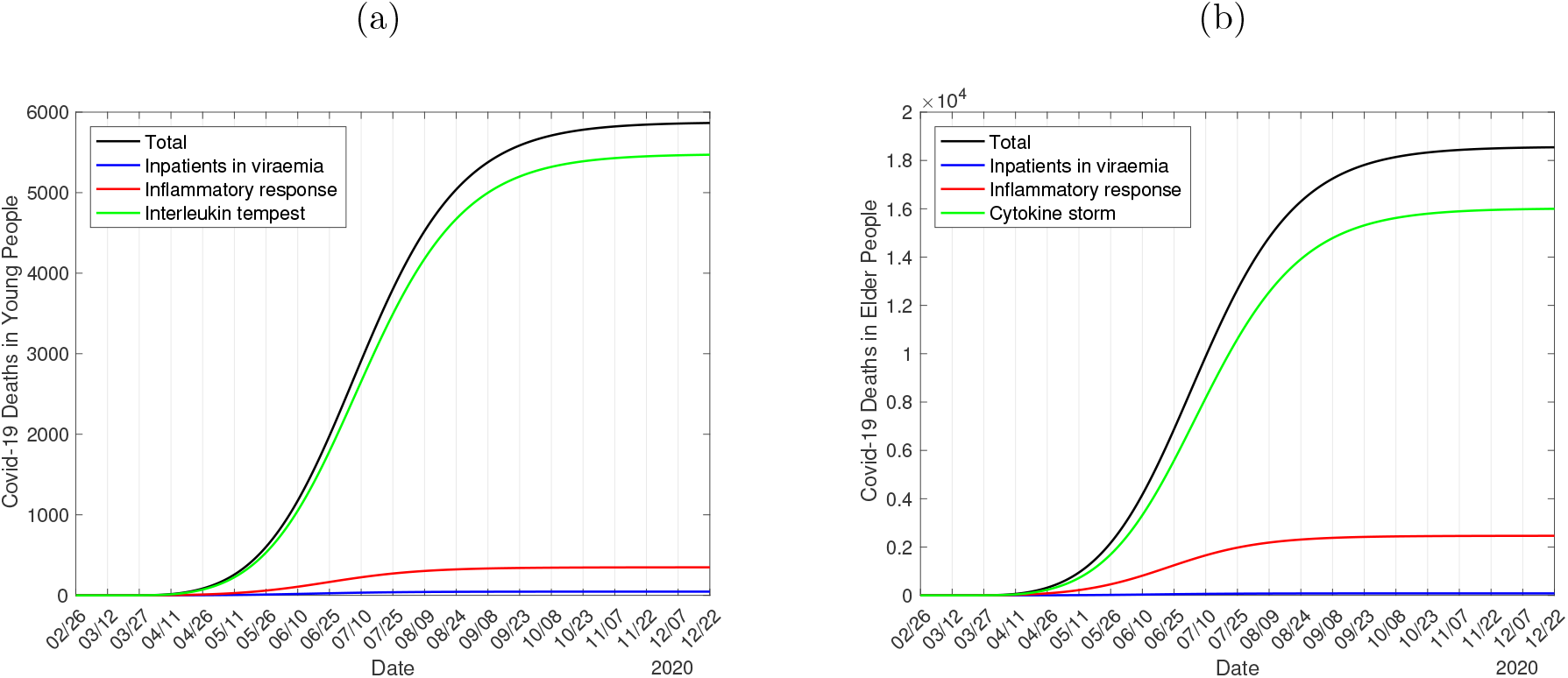
The calculated curves of П_1_, П_2_, П_3_, and П*_s_* = П_1_ + П_2_ + П_3_ for young (a) and elder (b) subpopulations.

In the natural epidemic, for young persons, the values of П_1_, П_2_, and П_3_, are 121, 908, and 14, 290, while for elder persons, the values are 205, 6,137, and 39, 870. The accumulated deaths for young, elder, and total persons are, respectively, 15, 319 (117%), 46, 212 (105%), and 61, 531 (107%). The percentage between parentheses is the ratio with respect to the deaths estimated from *D*_2_. Therefore, the estimated number of deaths from severe covid-19 cases is 7% lower than that estimated from the clinical evolution.

The current epidemiological scenario in São Paulo State is described by the values of model parameters given in Table II, and the values for the treatment of covid-19 in hospitals given in Table III. Especially the values given in Table III can vary as the protocols of treatment are improved, decreasing the number of deaths due to covid-19. However, we do not vary any values given in Tables II and III to describe the epidemiological scenarios of release.

### B. Epidemiological scenarios of release

To evaluate the epidemiological scenarios of release, we adapt the plan proposed by São Paulo State [5]: (a) phase 2 – 20% of capacity of each location, (b) phase 3 – 40% of capacity, (c) phase 4 – 60% of capacity, and (d) phase 5 – 100% of capacity. The progressive flexibilization depends on the epidemiological scenario, hence, there is not a fixed date to the progression. There is a possibility to return to the previous stage. Hence, we consider the scheme where the isolated population is divided into four equal releases, that is, 20% (*l*_1_*_j_* = 0.2), 40% (*l*_2_*_j_* = 0.25), 60% (*l*_3_*_j_* = 1/3), and 100% (*l*_4_*_j_* = 1) of the isolated population, for *j* = *y*, *o*. We also vary the protective factor *ε*.

We consider three strategies of release:

**Strategy A**.: 14 days between successive releases. The reason behind this strategy is the impact of release will be observed in the severe covid-19 cases around 9 days later.

**Strategy B**.: 21 days between successive releases. The reason behind this strategy is the impact of release will be observed in the deaths induced by severe covid-19 cases around 15 days later.

**Strategy C**.: The goal of this scheme of release is the protection of the elder subpopulation. Initially, we consider two regimes in strategies A and B:

**Regime 1**.: The first release on June 29 – In strategy A, the successive releases occur on July 13, 27, and August 10. In strategy B, the successive releases occur on July 20, August 10 and 31.

**Regime 2**.: The first release on July 13 – In strategy A, the successive releases occur on July 27, August 10 and 24. In strategy B, the successive releases occur on August 3, 24, and September 14.

From Figure 3, the peaks of the daily Ω*_d_* and severe cases *D*_2_ are, respectively, 4, 468 and 57, 900, occurring on June 14 and 25. Hence, all releases in regimes 1 and 2 occur after the peak (descending phase) of the epidemic.

From Figure 4(b), (1) on February 26 we have *R_ef_* = *R*_0_ = 9.24, (2) on March 24 (beginning of the isolation), *R_ef_* = 9.22 jumps down to *R_r_* = 4.35 given by equation (13), (3) on April 4 (adoption of the protective measures), *R_ef_* = 4.29 jumps down to *R_p_* = 2.15 given by equation (14), and (4) on May 1 (more contribution of the small cities, interiorization of the epidemic), *R_ef_* = 1.96 jumps down to *R_in_* = 1.78 given by equation (15). All these interventions decreased *R_ef_*, flattening the epidemic curve. However, the release of the isolated persons increases *R_ef_* resulting in the rebound of the epidemic. At the beginning of the release, in regime 1 on June 29, *R_ef_* = 0.788 jumps up to a higher value *R_ef_* = *R_u_* = 1.229, a gap of Δ*_u_* = 0.441, where the increased reproduction number *R_u_* is given by equation (16), and in regime 2 on July 13, *R_ef_* = 0.624 jumps up to a higher value *R_ef_* = *R_u_* = 1.065, a gap of Δ*_u_* = 0.441. And so on for the next releases.

#### 1. Strategy A – 14 days between releases

Figure 8 shows the curves of *D*_2_ with (continuous curve) and without (dashed curve) release for strategy A in regime 1 (a) and regime 2 (b). In regime 1, the first gap is Δ*_u_* = 0.441, which is quite the same for the next releases, except in the last release, when Δ*_u_* = 0.881. In the first three releases, 20% of the isolated population is released, while in the last release, the remaining 40%. In regime 2, the gaps are Δ*_u_* = 0.441 and Δ*_u_* = 0.88.

**FIG. 8:**
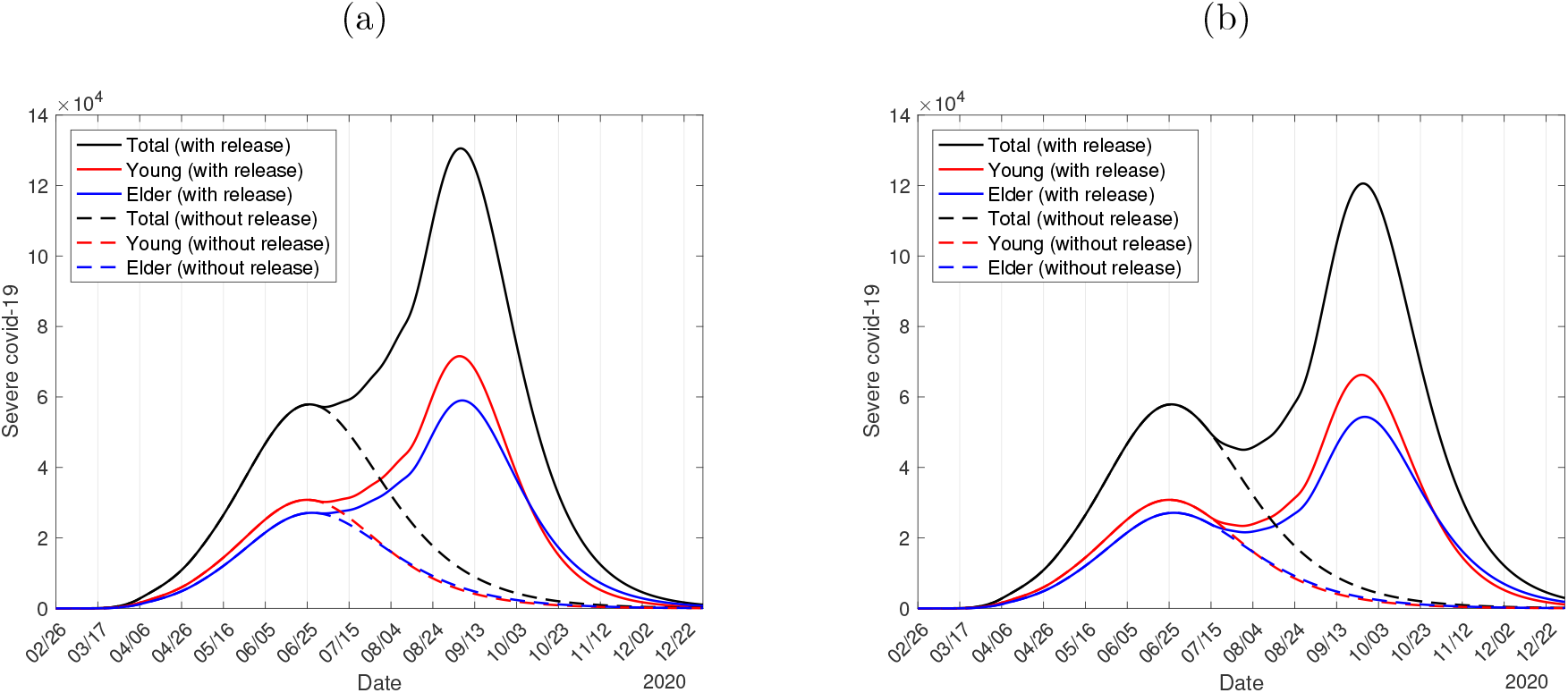
The curves of *D*_2_ with and without release for strategy A in regime 1 (a) and regime 2 (b).

Figure 8 shows the peak of the epidemic without release, see Figure 3(b), and the epidemic with release rebounds after this peak. For regime 1, the peaks of *D*_2_ for young and elder persons are, respectively, 71, 580 and 59, 010, occurring on September 5 and 6. For regime 2, the peaks of *D*_2_ for young and elder persons are, respectively, 66, 280 and 54, 360, occurring on September 25 and 26. For regime 1, the numbers of accumulated severe covid-19 cases for young, elder and total persons are, respectively, 575,900 (95.1%), 329,100 (96.5%), and 905, 000 (95.6%), and for regime 2, 573, 300 (94.7%), 327, 800 (96.1%), and 901,100 (95.2%). For regime 1, the numbers of accumulated immune young, elder and total persons are, respectively, 35.8 million (95.5%), 6.5 million (97%), and 42.3 million (95.7%), and for regime 2, 35.6 million (94.9%), 6.5 million (97%), and 42.1 million (95.2%). The percentage between parentheses is the ratio with respect to the natural epidemic.

In Figure 9, we show the calculated curves of *B*_2_, *B*_3_, and *B* = *B*_2_ + *B*_3_, for young (continuous curves) and elder (dashed curves) subpopulations for strategy A in regime 1 (a) and regime 2 (b). For regime 1, for young persons, the peaks of *B*_2_ and *B*_3_ are 6, 705 and 4, 426, occurring on September 6 and 16, while for elder persons, the peaks are 7, 666 and 8,635, occurring on September 5 and 15. For regime 2, for young persons, the peaks of *B*_2_ and *B*_3_ are 6, 231 and 4, 084, occurring on September 25 and October 6, while for elder persons, the peaks are 7,137 and 7, 978, occurring on September 23 and October 4.

**FIG. 9:**
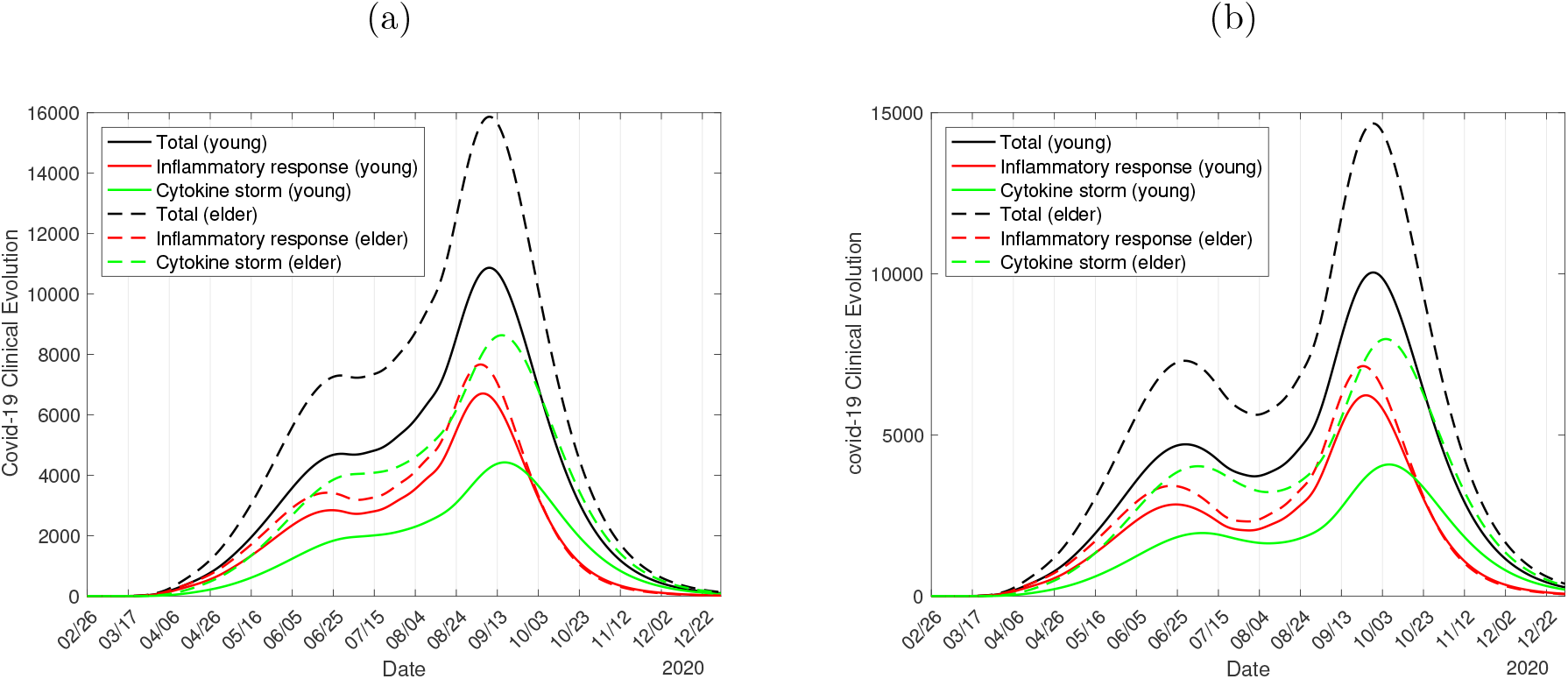
The calculated curves of *B*_2_, *B*_3_, and *B* = *B*_2_ + *B*_3_, for young (continuous curves) and elder (dashed curves) subpopulations for strategy A in regime 1 (a) and regime 2 (b).

The peaks of inpatients and ICUs patients occur very closely in young and elder subpopulations. For regime 1, on September 5, the number of inpatients is 14, 371 and on September 15, ICUs patients are 13, 061. For regime 2, on September 23, the number of inpatients is 13, 368 and on October 4, ICUs patients are 12, 062. The delay in 14 days to initiate the release resulted in a decrease of around 1, 000 patients in hospitals and ICUs and the peaks are delayed by 20 days.

The curves of cures and deaths are similar to those shown in Figures 6(b) and 7. At the end of the first wave of the epidemic, for regime 1, the numbers of accumulated cure for young (*C_y_*) and elder (*C_o_*) persons are, respectively, 558,100 and 282, 800, and the accumulated deaths П_1_, П_2_, and П_3_ for young persons are, respectively, 115, 863, and 13,560, while for elder persons, the values are 197, 5, 920, and 38, 380. The total numbers of deaths in young and elder subpopulations are, respectively, 14,538 (2.5%) and 44,497 (13.5%), totalizing 59, 035 (6.5%). For regime 2, the numbers of accumulated cure for *C_y_* and *C_o_* are, respectively, 555, 600, and 281, 700, and the accumulated deaths П_1_ П_2_, and П_3_ for young persons are, respectively, 115, 859, and 13, 430, while for elder persons, the values are 197, 5, 893, and 38, 080. The total numbers of deaths in young and elder subpopulations are, respectively, 14, 404 (2.5%) and 44,170 (13.5%), totalizing 58, 574 (6.5%). The delay in 14 days to initiate the release resulted in a decrease of 461 deaths. The percentage between parentheses is the severe covid-19 case fatality rate П/Ω. In the natural epidemic, the total number of deaths is 61, 531, and regimes 1 and 2 reduced the deaths in, respectively, 2, 496 (4.1%) and 2, 957 (4.8%).

To avoid the overloading in the health care system with the release, we assess how the protective measures and social distancing must be adopted and enhanced by the circulating population. Figure 10 shows, for *ε* = 0.3, the curves of *D*_2_ with (continuous curve) and without (dashed curve) release according to strategy A in regime 1 (a), and the calculated curves of *B*_2_, *B*_3_, and *B* = *B*_2_ + *B*_3_, for young (continuous curves) and elder (dashed curves) subpopulations (b). If we use *ε* = 0.33, we obtained quite the same results for regime 2.

**FIG. 10:**
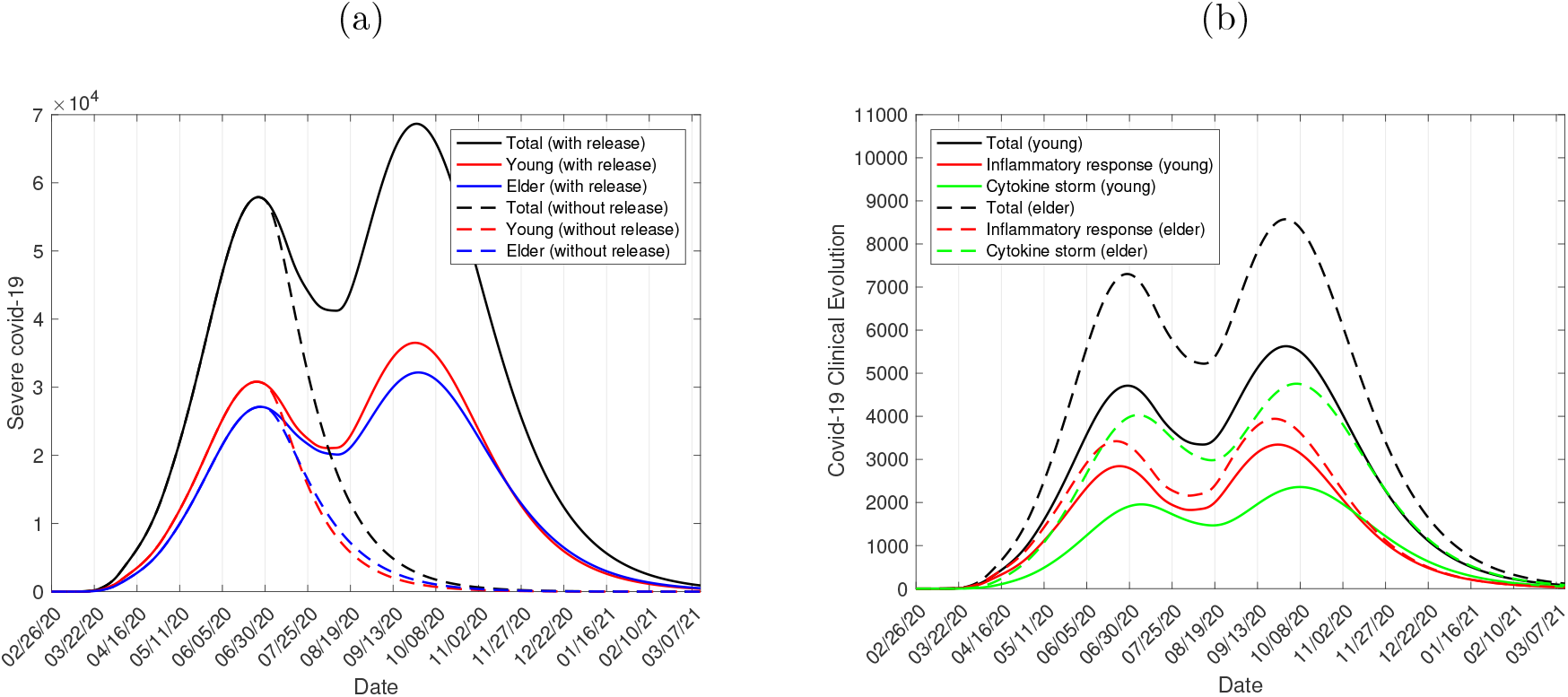
The curves of *D*_2_ with and without release for strategy A in regime 1 (a), and the calculated curves of *B*_2_, *B*_3_, and *B* = *B*_2_ + *B*_3_, for young (continuous curves) and elder (dashed curves) subpopulations (b) for *ε* = 0.3.

Figure 10 shows the peak of the epidemic without release, and the epidemic with release rebounds after this peak. The peaks of *D*_2_ for young and elder persons are, respectively, 36, 520 and 32,170, occurring on September 25 and 28. The peaks of *B*_2_ and *B*_3_ for young persons are 3, 344 and 2, 359, occurring on September 25 and October 7, while for elder persons, the peaks are 3, 942 and 4, 757, occurring on September 23 and October 5. The peaks of inpatients and ICUs patients occur very closely in young and elder subpopulations.

In strategy A, the need for ICUs increases continuously (regime 2 presents a little decreasing) up to the peak of occupancy, 13, 061 for regime 1, and 12,062 for regime 2. The capacity of ICUs in São Paulo State is 9,000, which could be increased. However, the persistently high number of inpatients and ICUs patients may lead to the exhaustion of health care professionals.

Notice that if the release had been initiated on June 15, only the first release in both regimes would occur in the ascending phase of the epidemic, and *R_ef_* = 1.022 jumped up to a higher value *R_ef_* = *R_u_* = 1.463, a gap of Δ*_u_* = 0.441. This strategy has results quite similar to those shown in Figures 8(a) and 9(a), but worse. For instance, the epidemic curve *D*_2_ increases without the small perturbation on June 25, and the peak is 135, 500, which occurs on August 30.

#### 2. Strategy B – 21 days between releases

Figure 11 shows the curves of *D*_2_ with (continuous curve) and without (dashed curve) release according to strategy B in regime 1 (a) and regime 2 (b). In regime 1, the first gap in jumping up is Δ*_u_* = 0.441, which is quite the same for the next releases, except in the last release, when Δ*_u_* = 0.88. In the first three releases, 20% of the isolated population is released, while in the last release, the remaining 40%. In regime 2, the gaps are Δ*_u_* = 0.441 and Δ*_u_* = 0.88.

**FIG. 11:**
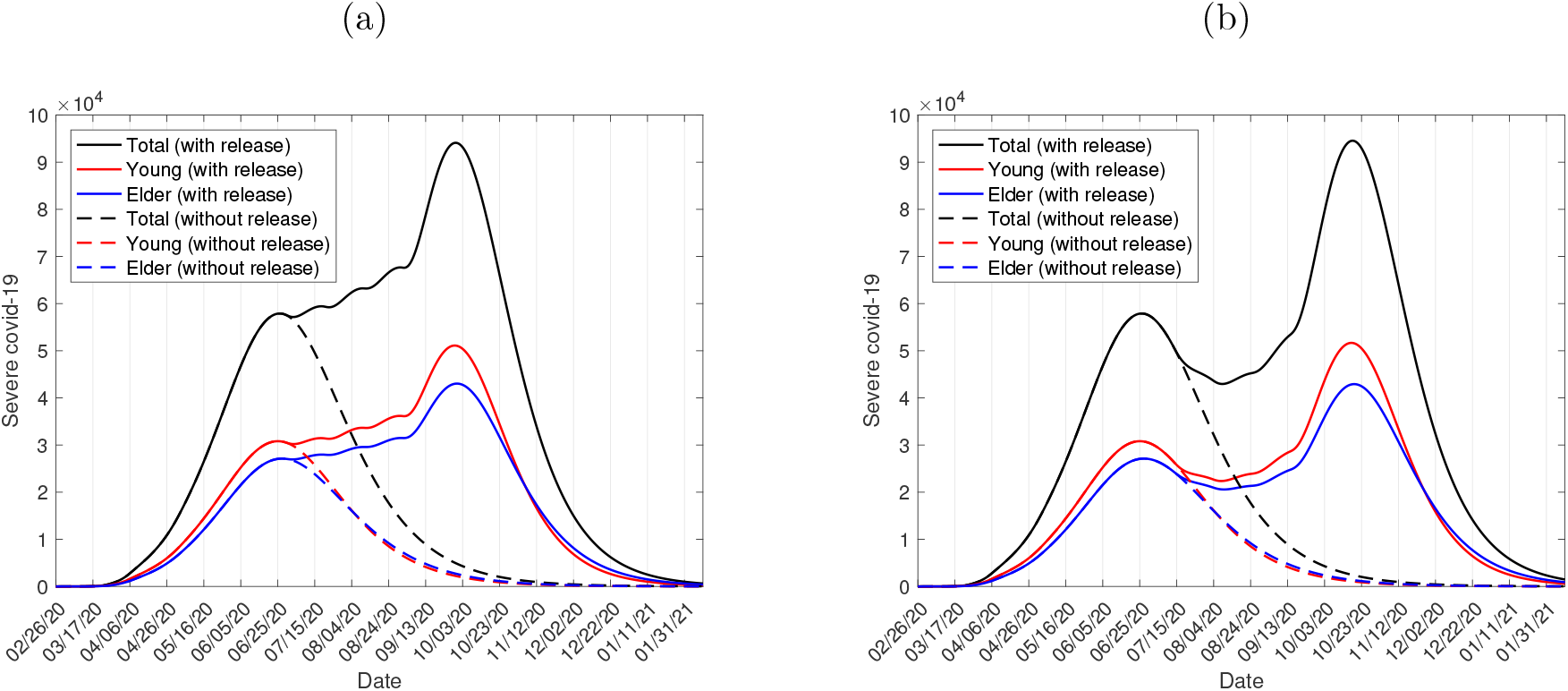
The curves of *D*_2_ with and without release for strategy B in regime 1 (a) and regime 2 (b).

Figure 11 shows the peak of the epidemic without release, and the epidemic with release rebounds after this peak. For regime 1, the peaks of *D*_2_ for young and elder persons are, respectively, 51,100 and 43, 030, occurring on September 28 and 29. For regime 2, the peaks of *D*_2_ for young and elder persons are, respectively, 51, 670 and 42, 920, occurring on October 17 and 18.

For regime 1, the numbers of accumulated severe covid-19 cases for young, elder and total persons are, respectively, 565,900 (93.5%), 324, 200 (95.1%), and 890,100 (94.1%), and for regime 2, 566,100 (93.5%), 324, 200 (95.1%), and 890, 040 (94.1%). For regime 1, the numbers of accumulated immune young, elder and total persons are, respectively, 35.1 million (93.6%), 6.4 million (95.5%), and 41.5 million (93.9%), and for regime 2, 35.1 million (93.6%), 6.4 million (95.5%), and 41.5 million (93.9%). The percentage between parentheses is the ratio with respect to the natural epidemic.

In Figure 12, we show the calculated curves of *B*_2_, *B*_3_, and *B* = *B*_2_ + *B*_3_, for young (continuous curves) and elder (dashed curves) subpopulations for strategy B in regime 1 (a) and regime 2 (b). For regime 1, for young persons, the peaks of *B*_2_ and *B*_3_ are 4, 730 and 3, 234, occurring on September 28 and October 9, while for elder persons, the peaks are 5, 405 and 6, 302, occurring on September 27 and October 8. For regime 2, for young persons, the peaks of *B*_2_ and *B*_3_ are 4,816 and 3, 234, occurring on October 17 and 28, while for elder persons, the peaks are 5, 509 and 6, 310, occurring on October 15 and 26.

**FIG. 12:**
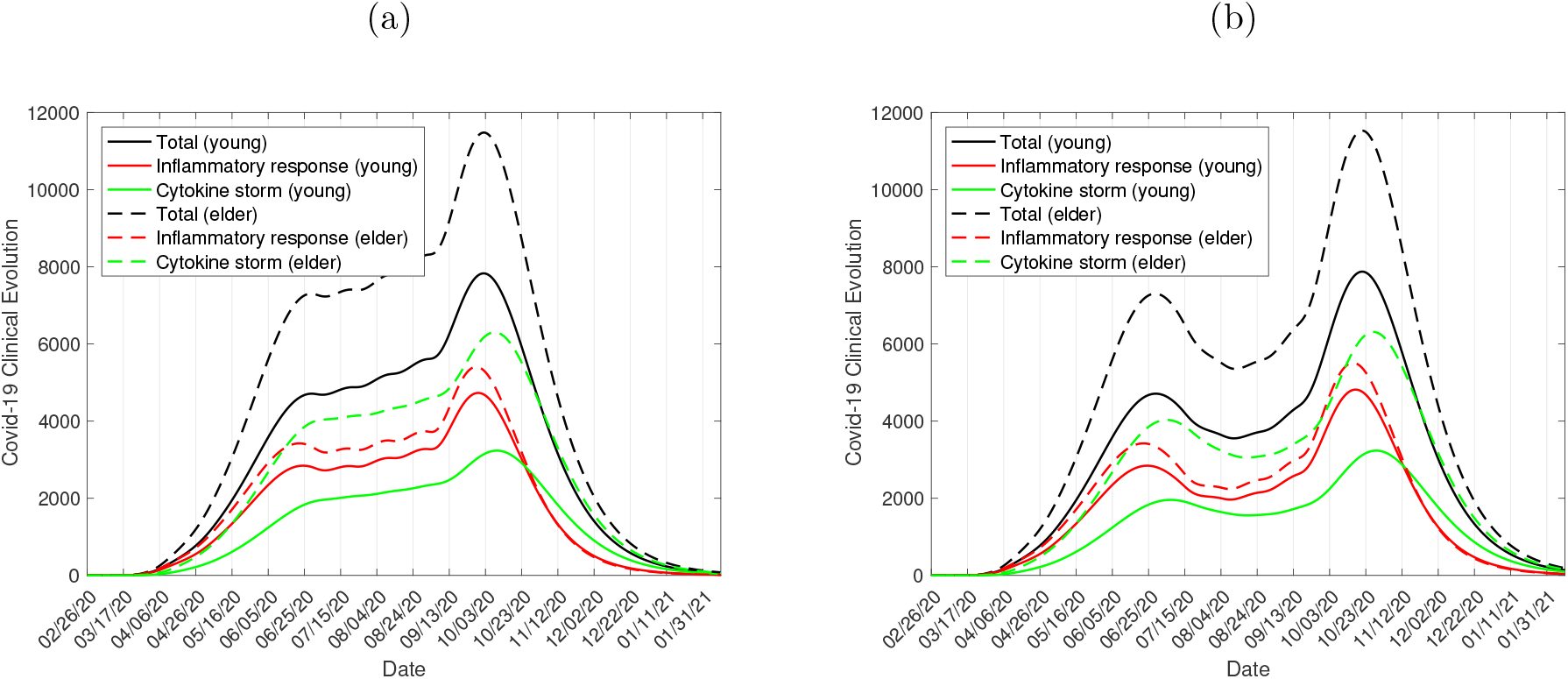
The calculated curves of *B*_2_, *B*_3_, and *B* = *B*_2_ + *B*_3_, for young (continuous curves) and elder (dashed curves) subpopulations for strategy B in regime 1 (a) and regime 2 (b).

The peaks of inpatients and ICUs patients occur very closely in young and elder subpopulations. For regime 1, on September 27, the number of inpatients is 10,135 and on October 8, ICUs patients are 9, 536. For regime 2, on October 15, the number of inpatients is 10, 325 and on October 26, ICUs patients are 9, 544. The delay in 14 days to initiate the release increased to around 200 patients in hospitals, and the peaks are delayed by 20 days. However, the hospital and ICUs care efforts in regime 1 is continuously increasing, which is much worse than in regime 2.

At the end of the first wave of the epidemic, for regime 1, the accumulated cures for young (*C_y_*) and elder (*C_o_*) persons are, respectively, 547,800 and 278, 300, and the accumulated deaths П_1_, П_2_, and П_3_ for young persons are, respectively, 113, 848, and 13, 340, while for elder persons, the values are 195, 5, 832, and 37, 850. The total numbers of deaths in young and elder subpopulations are, respectively, 14, 301 (2.5%) and 43, 877 (13.5%), totalizing 58,178 (6.5%). For regime 2, the accumulated cures for young (*C_y_*) and elder (*C_o_*) persons are, respectively, 548, 200 and 278, 400, and the accumulated deaths П_1_, П_2_, and П_3_ for young persons are, respectively, 113, 848, and 13, 310, while for elder persons, the values are 195, 5, 832, and 37, 780. The total numbers of deaths in young and elder subpopulations are, respectively, 14, 271 (2.5%) and 43, 807 (13.5%), totalizing 58, 078 (6.5%). The delay in 14 days to initiate the release resulted in a decrease of 100 deaths. The percentage between parentheses is the severe covid-19 case fatality rate П/Ω. In the natural epidemic, the total number of deaths is 61, 531, and regimes 1 and 2 reduced the deaths in, respectively, 3, 353 (5.4%) and 3, 453 (5.6%).

In strategy B, the need for ICUs increases continuously in regime 1 up to the peak of occupancy, 9, 536. As in strategy A, this continuous effort may lead to the exhaustion of health care professionals. However, for regime 2, the peak 9, 544 occurs 20 days later than regime 1 but presents a large period where the number of patients needing ICUs decreases, which may facilitate the work of health care professionals.

São Paulo State is planning to open schools on September 8, if at that date the previous releases of persons remain in yellow phase [5]. Notice that strategy B in regime 2 with the last release of 40% of remaining persons in isolation will be done on September 14, 6 days later. Hence, let us consider strategy B in regime 3, where on September 8 we have the third release:

**Regime 3.:** The first release on July 28 – In strategy B, the successive releases occur on August 18, September 8, and 29.

In strategy B in regime 3, at the beginning of the release on July 28, *R_ef_* = 0.516 jumps up to a higher value *R_ef_* = = 0.956, a gap of Δ*_u_* = 0.44, which is quite the same for next releases, except in the last release, when Δ*_u_* = 0.88.

The epidemic curve of *D*_2_, as well as the curves of *B*_2_ and *B*_3_ for strategy B in regime 3, are very similar to those shown in Figures 11(b) and 12(b), except the depression between two peaks is decreased by around, respectively, 10,000 and 2, 000. The peaks of *D*_2_ for young and elder persons are, respectively, 53, 530 and 43,980, occurring on November 6 and 8. The peaks of *B*_2_ and *B*_3_ for young persons are 5,010 and 3, 325, occurring on November 6 and 18, while for elder persons, the peaks are 5, 728 and 6, 487, occurring on November 5 and 16. The peaks of inpatients and ICUs patients occur very closely in young and elder subpopulations. On November 5, the patients in hospital care are 10, 738 and on November 16, the patients in ICUs care are 9, 812.

At the end of the first wave of the epidemic, for regime 3, the numbers of accumulated severe covid-19 cases for young, elder, and total persons are, respectively, 567, 600 (94%), 324, 800 (95%), and 892, 400 (94%). The numbers of accumulated immune young, elder, and total persons are, respectively, 35.2 million (93.9%), 6.4 million (95.5%), and 41.6 million (94.1%). The percentage between parentheses is the ratio with respect to the natural epidemic. The accumulated cure for young (*C_y_*) and elder (*C_o_*) persons are, respectively, 549, 300, and 278, 700, and the accumulated deaths П_1_, П_2_, and П_3_ for young persons are, respectively, 114, 850, and 13, 360, while for elder persons, the values are 195, 5, 843, and 37, 900. The total numbers of deaths in young and elder subpopulations are, respectively, 14, 324 (2.5%) and 43, 938 (13.5%), totalizing 58, 262 (6.5%). The percentage between parentheses is the severe covid-19 case fatality rate П/Ω. In the natural epidemic, the total number of deaths is 61, 531, and regime 3 reduced the deaths in 3, 269 (5.3%).

Hence, the benefit of the regime 3, which is regime 2 delaying the beginning of release in 15 days, is very little in comparison with regime 2.

#### 3. Strategy C - Protecting elder subpopulation

Figures 11(b) and 12(b), strategy B in regime 2, showed the best scenario for release. Let us consider strategy B and regime 2 for the release of the isolated young population, but all isolated elder persons are released later, called strategy C. The reason behind this strategy is the fact that elder subpopulation has 3-time more risk of infection when in interaction with young subpopulation.

In strategy C, the release of only young persons occurs on July 13, August 3, 24, and September 1. Hence, at the beginning of the release of young persons on July 13, *R_ef_* = 0.624 jumps up to a higher value *R_ef_* = = 0.993, a gap of Δ*_u_* = 0.369, which is quite the same for next two releases, except in the fourth releases, when Δ*_u_* = 0.736, and in the last release, when Δ*_u_* = 0.36. For the release of elder persons, we consider 3 regimes:

**Regime 4.:** Elder persons are released on September 28. At this day, *R_ef_* = 1.363 jumps up to a higher value *R_ef_* = *R_u_* = 1.723, a gap of Δ*_u_* = 0.33.

**Regime 5.:** Elder persons are released on December 1. At this day, *R_ef_* = 0.442 jumps up to a higher value *R_ef_* = *R_u_* = 0.801, a gap of Δ*_u_* = 0.359.

**Regime 6.:** Elder persons are released on January 1, 2021. At this day, *R_ef_* = 0.379 jumps up to a higher value *R_ef_* = = 0.738, a gap of Δ*_u_* = 0.359.

Figure 13 shows the curves of *D*_2_ with (continuous curve) and without (dashed curve) release according to strategy C in regime 4 (a), and the calculated curves of *B*_2_, *B*_3_, and *B* = *B*_2_ + *B*_3_, for young (continuous curves) and elder (dashed curves) subpopulations (b).

**FIG. 13:**
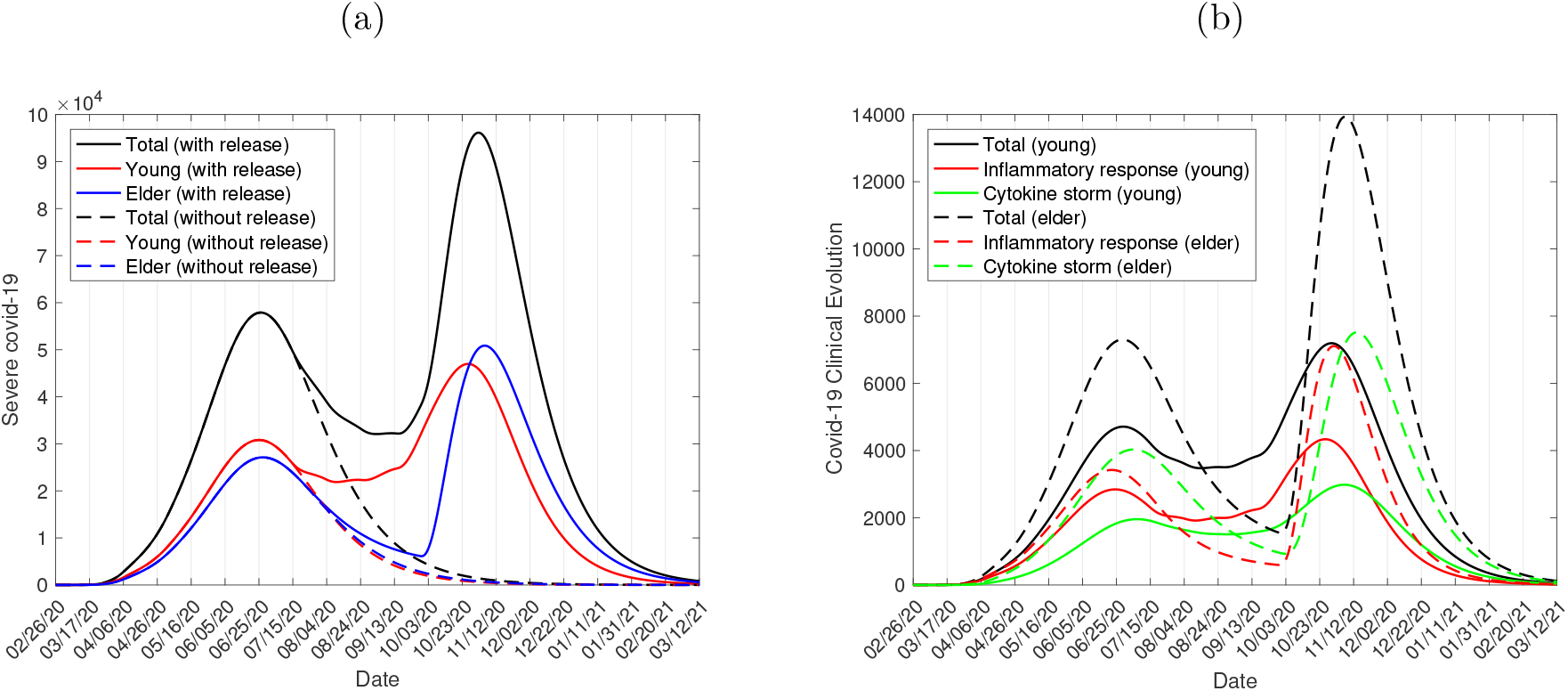
The curves of *D*_2_ with and without release strategy C in regime 4 (a), and the calculated curves of *B*_2_, *B*_3_, and *B* = *B*_2_ + *B*_3_, for young (continuous curves) and elder (dashed curves) subpopulations (b).

Figure 13 shows the peak of the epidemic without release, and the epidemic with release rebounds after this peak. The new peaks of *D*_2_ for young and elder persons are, respectively, 46, 980 and 50, 860, occurring on October 26 and November 5. The peaks of *B*_2_ and *B*_3_ for young persons are 4, 336 and 2, 980, occurring on October 26 and November 6, while for elder persons, the peaks are 7,109 and 7, 518, occurring on October 31 and November 14. The peaks of inpatients and ICUs patients occur very closely in young and elder subpopulations. On October 26, the number of inpatients is 11, 445 and on November 6, ICUs patients are 10,498.

For regime 4, the numbers of accumulated severe covid-19 cases for young, elder, and total persons are, respectively, 568, 700 (94%), 315, 700 (93%), and 884,400 (93%). The numbers of accumulated immune young, elder, and total persons are, respectively, 35.3 million (94.1%), 6.2 million (92.5%), and 41.5 million (93.9%). The percentage between parentheses is the ratio with respect to the natural epidemic.

At the end of the first wave of the epidemic, for regime 4, the accumulated cures for young (*C_y_*) and elder (*C_o_*) persons are, respectively, 550, 200 and 270, 900, and the accumulated deaths П_1_, П_2_, and П_3_ for young persons are, respectively, 114, 853, and 13,400, while for elder persons, the values are 190, 5, 679, and 36, 830. The total numbers of deaths in young and elder subpopulations are, respectively, 14, 367 (2.5%) and 42, 699 (13.5%), totalizing 57,066 (6.5%). The percentage between parentheses is the severe covid-19 case fatality rate n/Q. In the natural epidemic, the total number of deaths is 61, 531, and regime 4 reduced the deaths in 4, 465 (7.3%).

Figure 14 shows the curves of *D*_2_ with (continuous curve) and without (dashed curve) release according to strategy C in regime 5 (a), and the calculated curves of *B*_2_, *B*_3_, and *B* = *B*_2_ + *B*_3_, for young (continuous curves) and elder (dashed curves) subpopulations (b).

**FIG. 14:**
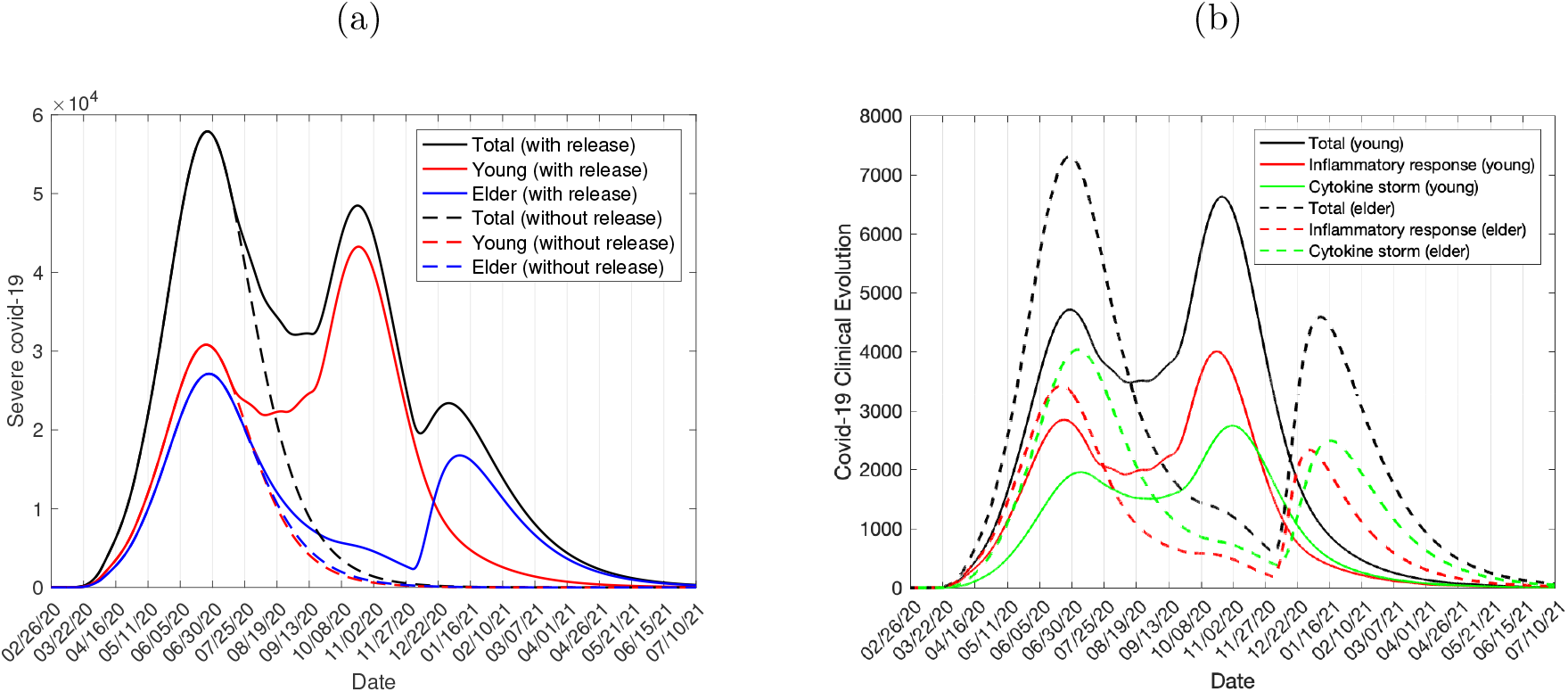
The curves of *D*_2_ with and without release strategy C in regime 5 (a), and the calculated curves of *B*_2_, *B*_3_, and *B* = *B*_2_ + *B*_3_, for young (continuous curves) and elder (dashes curves) subpopulation (b).

Figure 14 shows the peak of the epidemic without release, and the rebound of the epidemic but with a decreased second peak. The second peaks of *D*_2_ for young and elder persons are, respectively, 43, 260 and 16, 760, occurring on October 21 and January 7. The peaks of *B*_2_ and *B*_3_ for young persons are 4,003 and 2, 745, occurring on October 21 and November 1, while for elder persons, the second peaks are 2, 330 and 2, 493, occurring on January 1 and 16. The peaks of inpatients and ICUs patients occur very elapsed in young and elder subpopulations, facilitating the system health care.

For regime 5, the numbers of accumulated severe covid-19 cases for young, elder, and total persons are, respectively, 559, 300 (92%), 229, 800 (67%), and 789,100 (83%). The numbers of accumulated immune young, elder, and total persons are, respectively, 34.5 million (92%), 4.5 million (67.2%), and 39 million (88.2%). The percentage between parentheses is the ratio with respect to the natural epidemic.

At the end of the first wave of the epidemic, for regime 5, the accumulated cures for young (*C_y_*) and elder (*C_o_*) persons are, respectively, 539,900 and 196, 400, and the accumulated deaths П_1_, П_2_, and П_3_ for young persons are, respectively, 112, 839 and 13, 200 while for elder persons, the values are 138, 4,134, and 26, 830. The total numbers of deaths in young and elder subpopulations are, respectively, 14,151 (2.5%) and 31,102 (13.5%), totalizing 45, 253 (5.7%). The percentage between parentheses is the severe covid-19 case fatality rate П/Ω. In the natural epidemic, the total number of deaths is 61,531, and regime 5 reduced the deaths in 16, 278 (27%).

Figure 15 shows the curves of *D*2 with (continuous curve) and without (dashed curve) release according to strategy C in regime 6 (a), and the calculated curves of *B*_2_, *B*_3_, and *B* = *B*_2_ + *B*_3_, for young (continuous curves) and elder (dashed curves) subpopulations (b).

**FIG. 15:**
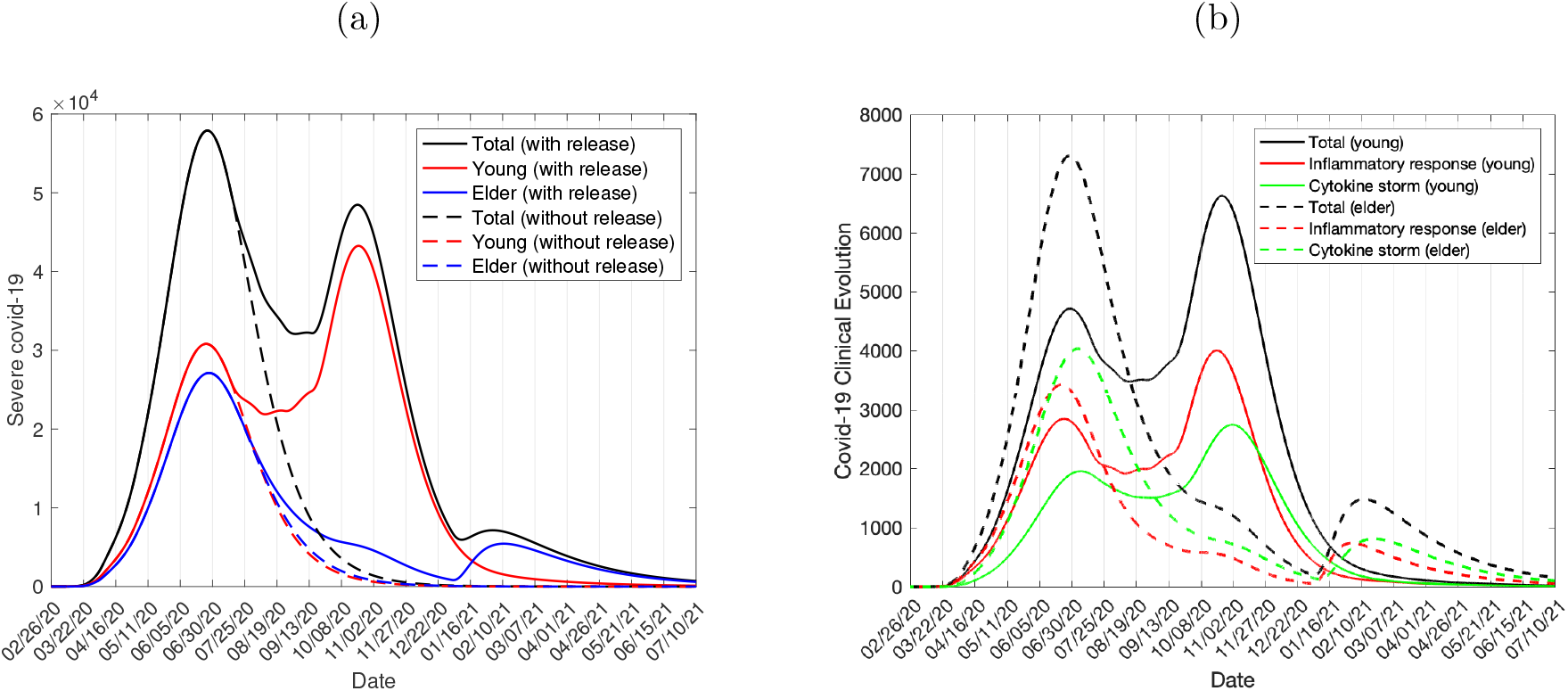
The curves of *D*_2_ with and without release strategy C in regime 6 (a), and the calculated curves of *B*_2_, *B*_3_, and *B* = *B*_2_ + *B*_3_, for young (continuous curves) and elder (dashed curves) subpopulations (b).

Figure 15 shows the peak of the epidemic without release, see Figure 3(b), and the rebound of the epidemic but with a decreased second peak. The second peaks of *D*_2_ for young and elder persons are, respectively, 43, 260 and 5, 444, occurring on October 21 and February 11. The peaks of *B*_2_ and *B*_3_ for young persons are 4, 003 and 2, 745, occurring on October 21 and November 1, while for elder persons, the second peaks are 740 and 812, occurring on February 3 and 20. The peaks of inpatients and ICUs patients occur very late in young and elder subpopulations, facilitating the health care system.

For regime 6, the numbers of accumulated severe covid-19 cases for young, elder, and total persons are, respectively, 552,800 (91%), 187,000 (55%), and 739,800 (78%). The numbers of accumulated immune young, elder, and total persons are, respectively, 34.1 million (90.9%), 3.7 million (55.2%), and 37.8 million (85.5%). The percentage between parentheses is the ratio with respect to the natural epidemic.

At the end of the first wave of the epidemic, for regime 6, the accumulated cures for young (*C_y_*) and elder (*C_o_*) persons are, respectively, 532, 600 and 159, 500, and the accumulated deaths П_1_, П_2_, and П_3_ for young persons are, respectively, 111, 829, and 13,040, while for elder persons, the values are 112, 3, 360, and 21, 760. The total numbers of deaths in young and elder subpopulations are, respectively, 13, 980 (2.5%) and 25, 232 (13.5%), totalizing 39, 212 (5.3%). The percentage between parentheses is the severe covid-19 case fatality rate П/Ω. In the natural epidemic, the total number of deaths is 61, 531, and regime 6 reduced the deaths in 22, 319 (36%).

In regimes 4-6, the elder population is released after the release of all young persons with the last release on September 1. The times of the release for elder persons are September 28 (regime 4), December 1 (regime 5) and January 1, 2021 (regime 6), remembering that the numbers of deaths in the natural epidemic for young, elder and total persons are, respectively, 15, 319, 46, 212, and 61, 531. For regime 4, the numbers of deaths are 14, 367 (94%), 42, 699 (92%), and 57, 066 (93%), for regime 5, 14,151 (92%), 31,102 (67%), and 45, 253 (74%), and for regime 6, 13, 980 (91%), 25, 232 (55%), and 39, 212 (64%). The percentage between parentheses is the ratio with respect to the natural epidemic. Notice that in regime 6, the number of deaths for elder persons is increased in 6, 685 to the epidemic without release (18, 547) in regime 6, but for young persons are increased in 8,115 to the epidemic without release (5, 865). Hence, the protection of elder persons benefited also young persons.

From Figure 13, the release of all elder persons by regime 4 occurs when the epidemic is in the ascending phase with the release of young persons, and the elder persons were not protected. At the end of the first wave of the epidemic, the epidemiological values are close to those in the natural epidemic. However, delaying the release of elder persons, the epidemic occurring among released young persons declines, and at the beginning of elder persons, the numbers of susceptible (consequently, *R_ef_*) and infected young persons are decreased. When the epidemic after the release of the young persons reaches lower values of *R_ef_* and infected persons, the released elder persons are not under increased risk of infection in comparison with a higher number of infected young persons. Hence, the delay in the release of elder persons may result in the epidemic occurring practically as isolated elder subpopulation. We pointed out that the epidemic in interaction with young persons increases strongly the risk of infection in elder persons (3-time), and the reproduction number for young and elder subpopulations are 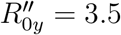 and 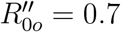.

Regimes 4-6 in strategy C showed that the decreased interaction by isolation between young and elder subpopulations protected the vulnerable elder persons. Hence, these regimes indicate that the number of deaths can be minimized if elder subpopulation and young persons with comorbidities (obesity, cardiopathy, diabetes, etc.) related to covid-19 could be protected. This protection can be isolation, and by vaccine when available.

From Figure 4(b), at the end of the first wave of the epidemic, we have *R_ef_* = 0.39. Figure 15(a) showed strategy of release C in regime 6 protecting the elder persons. Let us consider the isolation lasting until December 31, 2020, and on January 1, 2021, the isolated population is released, that is, all young and elder persons in classes *Q_y_* and *Q_o_* are transferred to *S_y_* and *S_o_*, resulting in

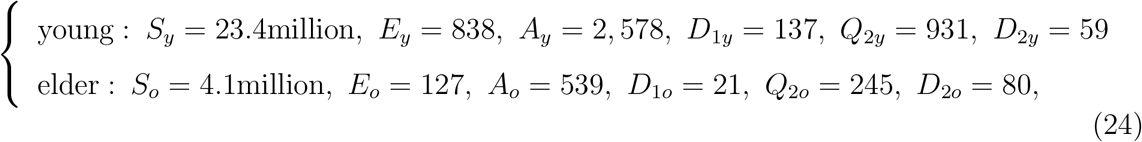

with *Q_y_* = *Q_o_* = *Q*_1_*_y_* = *Q*_1_*_o_* = *Q*_3_*_y_* = *Q*_3_*_o_* = 0 and *I* = 17.1 million. Hence, we consider two regimes:

**Regime 7.:** The first release on January 1, 2021 – The successive release of equal young and elders occur on January 22, February 12, and March 5. On January 1, 2021 (first release), *R_ef_* = 0.396 jumps up to a higher value *R_ef_* = *R_u_* = 0.834, a gap of ∆_u_ = 0.438, and on March 5 (last release), *R_ef_* = 1.714 jumps up to a higher value *R_ef_* = *R_u_* = 2.588, a gap of ∆*_u_* = 0.874.

**Regime 8.:** The first release on January 1, 2021 - All young and elder persons are released on January 1, 2021. On January 1, *R_ef_* = 0.396 jumps up to a higher value *R_ef_* = *R_u_* = 2.588, a gap of ∆*_u_* = 2.192.

The number of persons in each class given by equation (24) portrays regime 8. However, these values are practically the same for regime 7 on March 5, 2021.

Figure 16 shows the curves, for the release according to strategy C in regime 7 (dashed curve) and regime 8 (continuous curve), of *D*_2_ (a), and S (b).

**FIG. 16:**
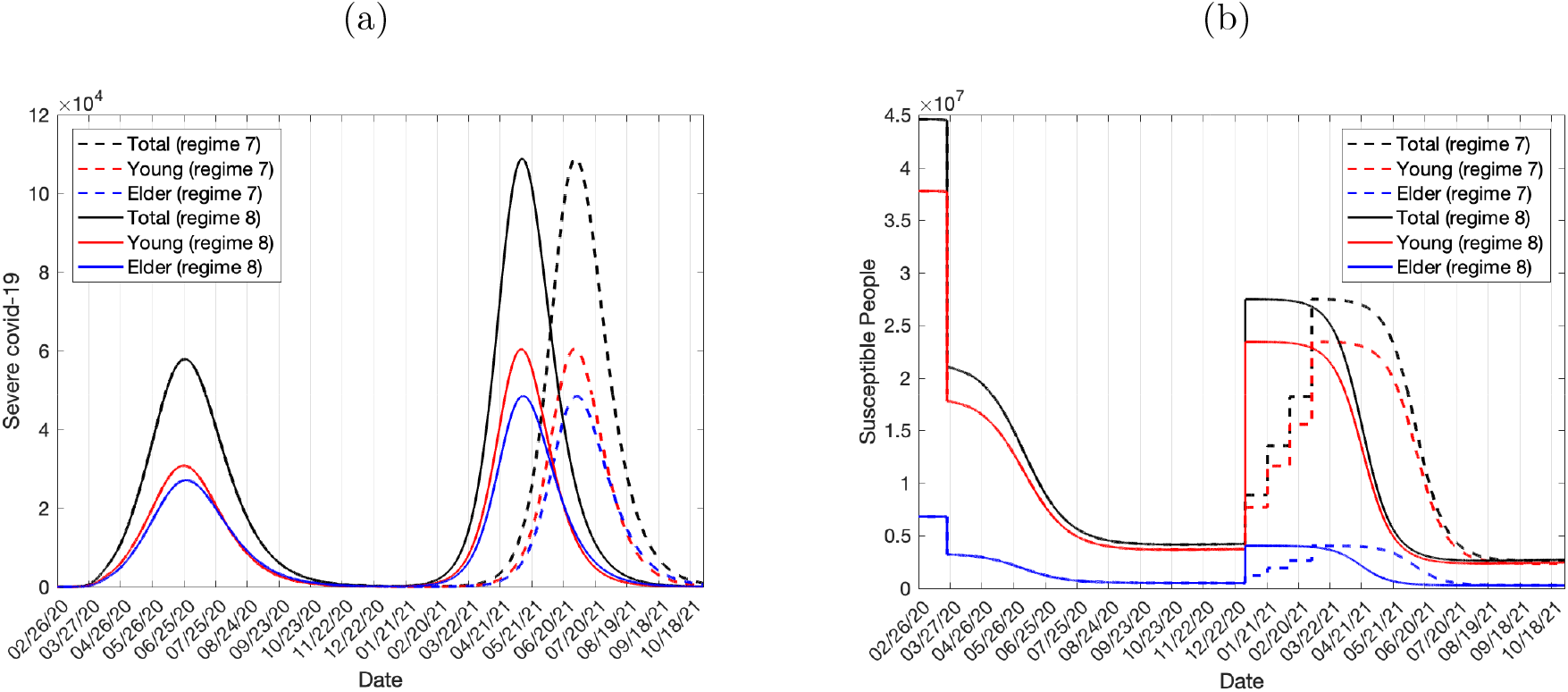
The curves for strategy C in regime 7 (dashed curve), and regime 8 (continuous curve) of *D*_2_ (a), and *S* (b).

Figure 16 shows the peak of the retaking epidemic after release. For regime 7, the retaken peaks of *D*_2_ for young and elder persons are, respectively, 60, 510 and 48,470, occurring on July 1 and 2, 2021. For regime 8, the peaks of *D*_2_ for young and elder persons are, respectively, 60,400 and 48, 520, occurring on May 11 and 12, 2021. At the end of the retaken wave of the epidemic, that is, end of the first wave of the epidemic, on October 31, 2021, for regime 7, the numbers of accumulated severe covid-19 cases for young, elder and total persons are, respectively, 573,900 (95%), 325,800 (96%), and 899, 700 (95%). The numbers of accumulated immune young, elder, and total persons are, respectively, 35.5 million (94.7%), 6.4 million (95.5%), and 41.9 million (94.8%). Quite the same values are obtained for regime 8. The percentage between parentheses is the ratio with respect to the natural epidemic.

At the end of the first wave of the epidemic, on October 31, 2021, for regime 7, the accumulated deaths П_1_, П_2_, and П_3_ for young persons are, respectively, 115, 860 and 13, 510, while for elder persons, the values are 195, 5,860, and 38,010. The total numbers of deaths in young and elder subpopulations are, respectively, 14,485 (2.5%) and 44,065 (13.5%), totalizing 58,550 (6.5%). The percentage between parentheses is the severe covid-19 case fatality rate П/Ω. In the natural epidemic, the total number of deaths is 61, 531, and regime 7 reduced the deaths in 2, 981 (4.8%). Quite the same values are obtained for regime 8. These regimes are better strategies of release to control the epidemic in the absence of effective treatment and vaccine, but hard to be implemented. Notice that all asymptotic values obtained in the preceding section for the epidemic under isolation correspond to those values at the time of the first release in regimes 7 and 8. Hence, the number of deaths during the retaken of the epidemic is 58, 550 − 24, 412 = 34,138.

At the moment when all persons are released in regimes 7 and 8, *R_ef_* = 0.396 jumps up to *R_ef_* = 2.588, but the number of infected individuals in circulation is very lower according to equation (24). Although higher value for *R_ef_* the reduced number of infected persons delayed the quick increasing phase of the forthcoming epidemic. In regime 7 (four releases elapsed in 21 days) the retaken of the epidemic occurred in around 4 months after the release. However, in regime 8 (unique release), the outbreak occurred in around 2 months after the release. In [4], we showed that, at the time of the beginning of release in several phases in Spain, the value of *R_ef_* was small, as well as the number of infected persons. Hence, regime 7 could explain the release strategy adopted in Spain. However, as shown in Figure 16, if rigid protective measures and the mass tests to isolated infected persons should not be continued, then the dammed epidemic can be released and trigger a severe outbreak.

## IV. DISCUSSION

We established the epidemiological scenario of isolation in São Paulo State by obtaining the flattened epidemic curve *D*_2_ and the effective reproduction number *R_ef_* during the epidemic. However, the basic reproduction number *R*_0_ = 9.24 due to successive control interventions decreases to 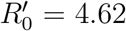 by the adoption of protective measures, and 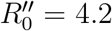 by the interiorization of the epidemic. Hence, 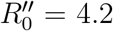 is the actual reproduction number when the release begins. From the epidemic curve, we retrieved the number of patients in the three phases of the clinical evolution to assess the occupancy of hospitals. This scenario was the background to study the epidemiological scenarios of release.

We analyzed three strategies of release. The flattened epidemic with interventions (isolation, social distancing, protective measures, and interiorization) was perturbed by the release of isolated persons. This perturbed epidemic is the continuation of the epidemic under isolation, and the entire epidemic curve forms the first wave of the epidemic. Depending on the scheme of release, the epidemic with the release was a rebound of the epidemic in the course or separated by a depression.

All strategies of release were compared with the natural epidemic. At the end of the first wave of the epidemic, strategies of release A and B in regimes 1-3 and strategy C in regime 4 reduced the numbers of accumulated severe covid-19 cases, immune persons, and deaths in around 5%. In other words, the susceptible persons are extremely reduced in these strategies of release. However, strategy C in regime 5 reduced the numbers of accumulated severe covid-19 cases and immune persons in around 15% and deaths in around 27%, while for regime 6, 18% and 36%. These two regimes fulfilled the goal of protecting elder persons. However, the relatively higher number of susceptible persons at the end of the first wave of the epidemic will originate a second wave before the second outbreak in the natural epidemic. Indeed, the second wave of the covid-19 natural epidemic in São Paulo State will occur in 2039, while for strategy C in regime 6, in 2036. When the first wave of the epidemic ends, the numbers of susceptible and infectious persons are very small, and the next second wave occurs only when the number of susceptible persons surpasses a threshold. However, the number of the infected persons can drop to zero due to the stochastic fluctuation, and the SARS-CoV-2 does not circulate anymore.

In all strategies and regimes, the epidemiological values achieved practically those values found in the natural epidemic. This finding is expected when the entire first wave of the epidemic is taken into account, once in São Paulo State, the actual reproduction number was 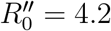 when the release began. From [4], at the end of the first wave of the epidemic, the accumulated severe covid-19 cases Ω for *R*_0_ = 9.24, 6.99, and 5.09 were very close, respectively, 946, 400, 945, 700, and 941, 500, however, the period of the first wave of the epidemic was enlarged as *R*_0_ decreased. For *R*_0_ > 4, the number of immune persons is quite that found in the natural epidemic.

The effective reproduction number *R_ef_* oscillates around one [10] and, at the end of the first wave of the epidemic, as shown in Figure 4(a), assumes lower value, which is lowered as *R*_0_ increases. Hence, disregarding the number of immune persons, the infections occur whenever susceptible persons (*R_ef_* depends on them) interact with infected persons. Herd immunity is a protection by vaccine aiming at the eradication of infections in a long-term epidemic, and in the short-term epidemic, the concept of herd immunity is meaningless (see discussion of herd immunity in [4]). In an epidemic triggered by an infection with the estimated basic reproduction number being *R*_0_, the critical proportion to be vaccinated is *p_c_* = 1 − 1/*R*_0_ [6], and if *p* > *p_c_*, the epidemic fades out.

In regimes 1-6, the release of isolated persons originated the rebound of the epidemic, however, Figure 16 showed the retaken of the epidemic after elapsed 2 (regime 8) and 4 (regime 7) months after the release. Hence, the release according to strategy C in regime 8 can be improved by enhancing the protective measures and sanitation and mass testing in the circulating population [11] to suppress the forthcoming epidemic. Figure 17 illustrates the rigid adoption of protective measures and social distancing, such that the protection factor is decreased to *ε* = 0.25 (continuous curve) and 0.3 (dashed curve) (a), and the isolation of especially those who are transmitting SARS-CoV-2 caught by mass testing, letting *η_j_* = *η*_1_*_j_* = *η*_2_*_j_* = 0.1 (continuous curve) and 0.05 (dashed curve) days^−1^, for *j* = *y*, *o*, but maintaining *ε* = 0.5 (b). However, both control mechanisms can be applied jointly to achieve better control of the covid-19 epidemic.

The epidemic of influenza pandemic in 1918 and 1919 occurred in London showed two peaks separated by 2 months [12], similar to 17(a). From Figure 17(a), when *ε* = 0.25, the peak of *D*_2_ is 15, 670 occurring on February 6, 2022, while for *ε* = 0.3, the peak of *D*_2_ is 36, 260 occurring on September 24, 2021. The reduced basic reproduction number 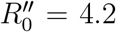 (partials 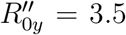 and 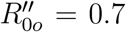) is reduced to 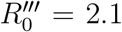 (partials 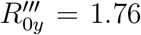 and 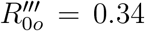) for *ε* = 0.25, and 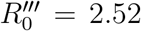 (partials 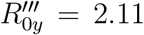 and 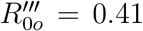) for *ε* = 0.3. For *ε* = 0.25, the numbers of accumulated severe covid-19 cases and deaths are, respectively, 637,840 and 39, 335, while for *ε* = 0.3, 746, 760 and 45, 930. In the natural epidemic, the total number of deaths 57, 340 (deaths from *D*_2_, Figure 2), which is reduced in 18,005 (31%) for *ε* = 0.25, and 11,410 (20%) for *ε* = 0.3.

**FIG. 17:**
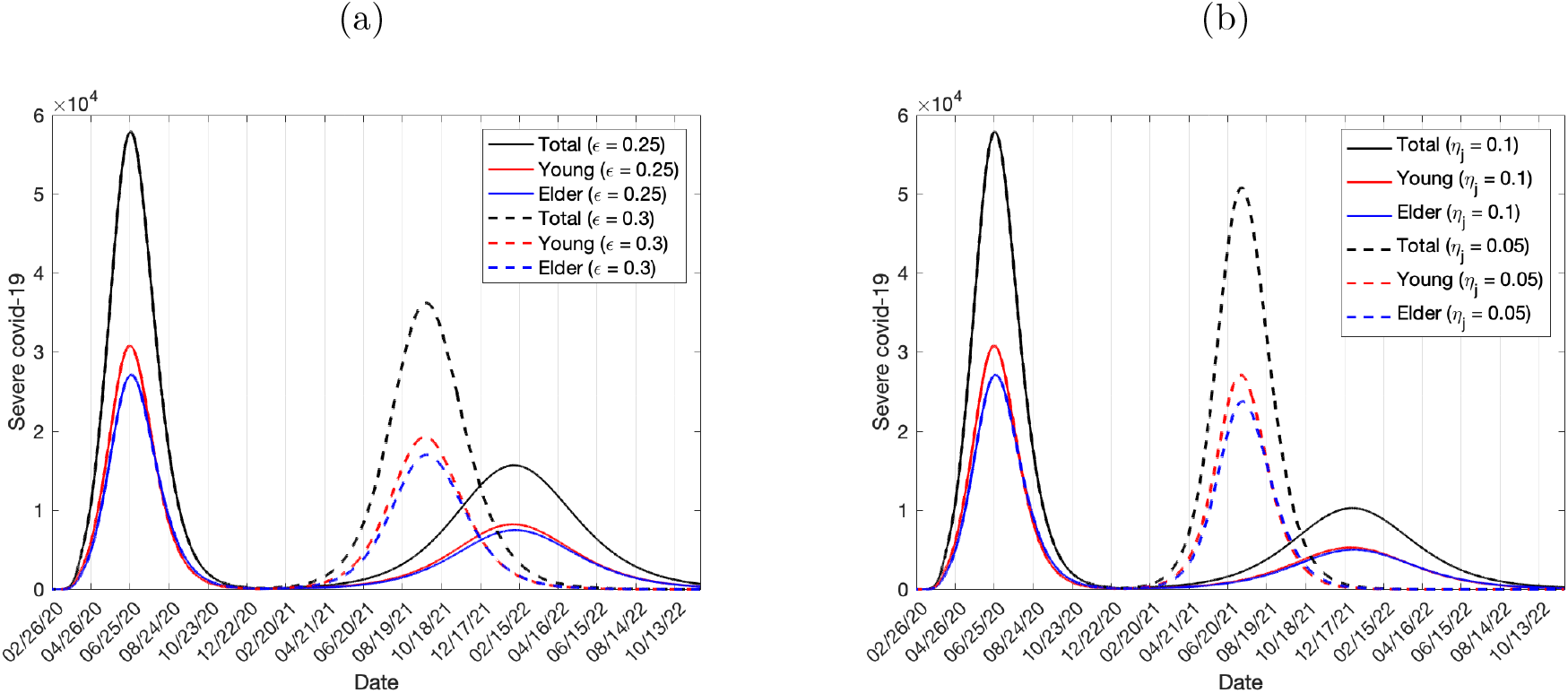
Illustration of the increased protective measures to *ε* = 0.25 (continuous curve) and 0.3 (dashed curve) (a), and the isolation by mass testing, letting *η_j_* = 0.1 (continuous curve) and 0.05 (dashed curve) *days*^−1^, for *j* = *y*, *o*, but maintaining *ε* = 0.5 (b).

From Figure 17(b), when *η_j_* = 0.1, the peak of *D*_2_ is 10, 250 occurring on December 29, 2021, while for *η_j_* = 0.05, the peak of *D*_2_ is 50,850 occurring on July 11, 2021. From equation (B8), the reduced basic reproduction number 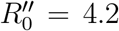 (partials 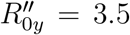 and 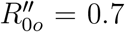) is reduced to 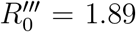 (partials 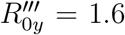 and 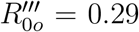) for *n_j_* = 0.1, and 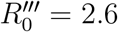 (partials 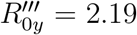 and 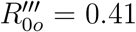) for *n_j_* = 0.05. For *n_j_* = 0.1, the numbers of accumulated severe covid-19 cases and deaths are, respectively, 545,090 and 33, 830, while for *η_j_* = 0.05, 746,820 and 46, 070. In comparison with the natural epidemic, the total number of deaths is reduced in 23, 510 (41%) for *η_j_* = 0.1, and 11, 270 (20%) for *η_j_* = 0.05.

As we have pointed out, on June 16 the curve of Ω detached from the epidemic in the course and presented increased trend (Figure 1(a)), as well as the curve of the accumulated deaths П on June 30 (Figure 2(a)). Hence, on June 7 and June 15 external factors resulted in the increased transmission of SARS-CoV-2. Let one of the external factors be the release of isolated persons. We evaluate the proportions of release on June 7 and 15 by estimating the accumulated deaths П, and the severe covid-19 cases Ω. Figure 18 shows the estimated curve of П (a), and the corresponding curve of Ω (b), using the proportions of estimated release *l* = 0.05 on June 7, and *l* = 0.05 on June 15.

**FIG. 18:**
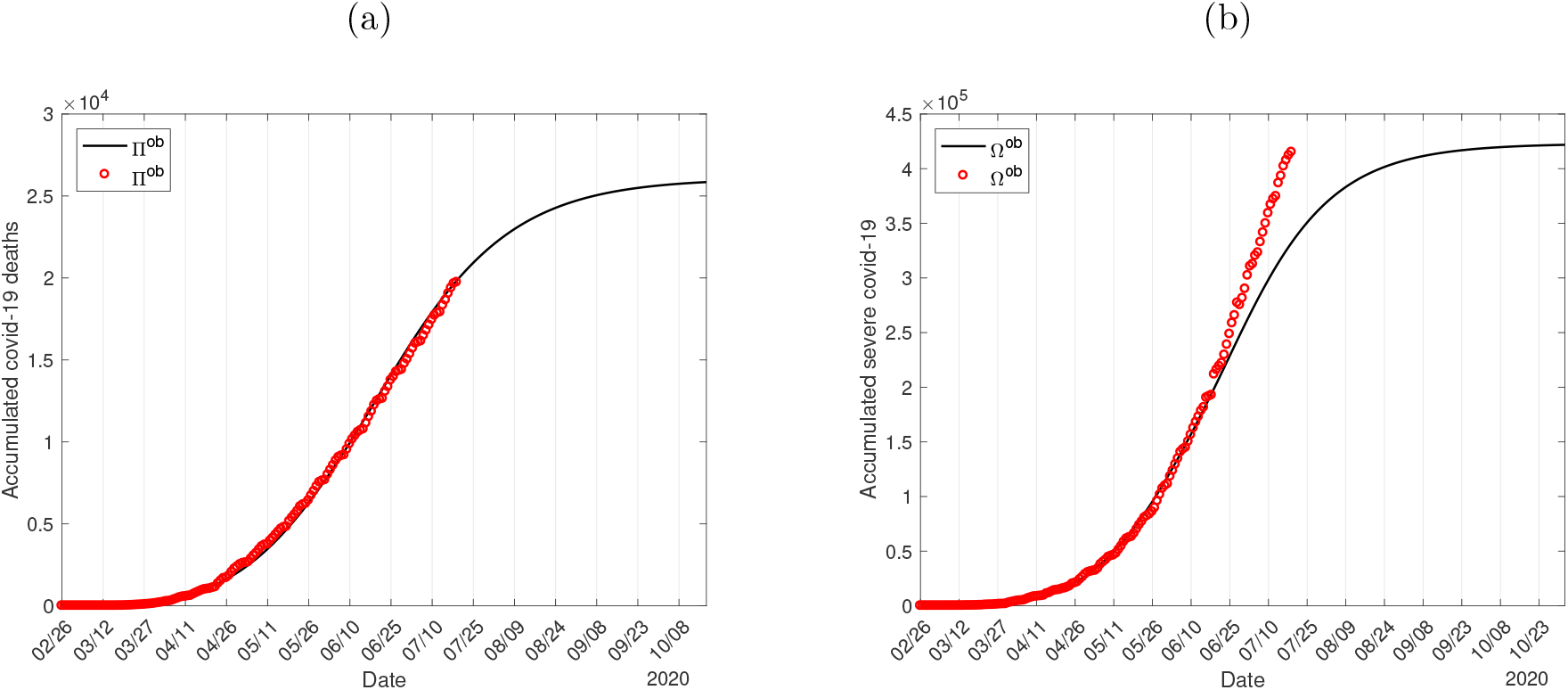
The estimated curve of П (a), and the curve of Ω (b), using the proportions of estimated release *l* = 0.05 on June 7, and *l* = 0.05 on June 15.

Daily covid-19 cases collected from São Paulo State contain both severe and mild cases, but covid-19 deaths are caused only by severe cases. All deaths registered in São Paulo State data collection were confirmed by the covid-19 test, hence these data are reliable. For this reason, we used daily registered deaths to estimate the curve of the number of deaths П, and the accumulated cases of covid-19 Ω did not match with the observed data. From Figure 18(b), the observed data situate above the curve of Ω, which confirms that the release of 10% of the isolated population can explain the new increase trend of the epidemic. Notice that the accumulated number of deaths was estimated until July 18, indicating that the curve of Ω under release is reliable only from June 16 to July 3 (15 days before the last recorded date of death), and the surplus number of registered covid-19 data may probably be due to the increased test of mild cases. However, since July 4, the additional number of covid-19 cases beyond the estimated curve of Ω may be originated by the mass test of mild covid-19 cases and/or increased release of the isolated persons.

In all strategies, the number of deaths was calculated assuming only the fatality due to the covid-19. However, if the severe covid-19 cases surpass the capacity of hospital care, then untreated persons could die, increasing the number of deaths beyond the fatality of covid-19. For this reason, the knowledge of the number of inpatients and, especially, the number of ICUs patients must be taken into account to choose a suitable strategy of release. In the strategies analyzed here, strategy C in regimes 5 (Figure 14), 6 (Figure 15), and 8 modified (Figure 17) avoided the overloading in hospitals. Notice that the clinical evolution parameters given in Table III can vary as the protocols of treatment are improved. When an effective and safe vaccine should be available, the model proposed here can be applied to evaluate the mass vaccination (changing the testing rates by vaccination rates), and Figure 17(b) can be interpreted as the impact of mass vaccination.

All results presented here were based on the homogeneity in time (model parameters do not vary), space (demographic density and social interaction do not vary) and individuals (age, gender, genetic, social condition), and also, in the absence of environmental fluctuation (seasonality) and stochasticity (small-sized populations), migratory movements (intense air traffic), and interventions (vaccination). These heterogeneities can be incorporated in more elaborated models, such as the age-structured transmission rate (see [13] and [14]), time-varying model parameters (see [15]), and stochastic parameter (see [16]).

For instance, let us illustrate the effect of demographic density in the covid-19 epidemic. The basic reproduction number *R*_0_ estimated for the covid-19 epidemic was 9.24 in São Paulo State, and 8.0 in Spain [4]. São Paulo State has 44.6 million inhabitants [17] with demographic density 177/*km*^2^ [18], while Spain has 47.4 million inhabitants [19] with demographic density 92.3/*km*^2^ [18]. The size of both populations is very close, but the demographic density in São Paulo State is around 2-time that in Spain. At the beginning of the epidemic, the daily registered collection of covid-19 data from a region (cities, states, and countries) is strongly influenced by the populational density, social contacts, etc. Hence, these features are taken into account by the estimated transmission rates *β_y_* and *β_o_* in the natural epidemic phase.

## V. CONCLUSION

We formulated a mathematical model considering two subpopulations composed of young and elder persons to study the covid-19 epidemic in São Paulo State, Brazil. The model considering pulses in isolation and releases was simulated to describe the epidemic under isolation in São Paulo State, and further epidemiological scenarios of release.

The importance of mathematical modeling is the possibility of evaluating different strategies of release to help public health authorities making decisions. We analyzed three strategies in eight regimes based on the plan proposed by the São Paulo State authorities. The epidemic curve under the release provided the estimation in the number of deaths and occupancy in hospitals and ICUs. All strategies showed that the number of deaths due to covid-19 was practically the same as that found in the natural epidemic. Hence, the number of deaths must not be a key parameter to plan release strategies. Rather, strategies of release must rely on the capacity of the health care system to treat adequately all severe covid-19 cases, which is proportional to the epidemic curve. Better strategies of release are those that the maximum occupancy of hospitals and the capacity of health care workers to treat patients during the epidemic must not be surpassed. This epidemic situation can be achieved when the beginning of several releases is delayed, which enlarges the period of the epidemic and avoids the collapse of the health care system.

The fatality rate of severe covid-19 among elder persons is higher than young persons. Additionally, the risk of elder persons being infected increases 3-time when in interaction with young persons. Hence, the isolation to control the covid-19 epidemic to some extent protected elder persons, and strategies of release must take this aspect into account. The better scheme aiming at the protection of elder persons is their release when the epidemic after the released young persons has reached a lower level. This strategy, however, is hard to be implemented, which is the reason to adopt lockdown or isolation.

## Data Availability

Data collected from São Paulo State government.

https://www.seade.gov.br/coronavirus/

## AUTHOR CONTRIBUTIONS

H. M.Y., L.P.L.J., A.C.Y. and F.F.M.C. contributed equally to this work.

## Appendix A: Model parameters, initial and boundary conditions

The system of equations (3), (4), and (5) in the main text is formulated based on the flowchart of the new coronavirus transmission model shown in Figure 19. The subscript *y* and *o* stand for, respectively, young and elder subpopulations.

From Figure 19, susceptible young persons in class *S_y_* are transferred to the class of susceptible elder persons *S_o_*, by the aging rate *φ*. For each subpopulation *j*, *j* = *y*, *o*, susceptible persons in class *S_j_* are infected by the force of infection λ*_j_* and enter into class *E_j_*. After an average period 1/*σ_j_* in class *E_j_*, where *σ_j_* is the incubation rate, exposed persons enter into the asymptomatic class *A_j_* (with probability *p_j_*) or pre-diseased class *D*_1_*_j_* (with probability 1 − *p_j_*). After an average period 1/*γ_j_* in class *A_j_*, where *γ_j_* is the recovery rate of asymptomatic persons, symptomatic persons acquire immunity (recovered) and enter into immune class *I*. Possibly asymptomatic persons can manifest symptoms at the end of this period, and a fraction 1 − *χj* enters into mild covid-19 class *Q*_2_*_j_*. Another route of exit from class *A_j_* is being caught by a test at a rate *η_j_* and enter into class *I* (we assume that this person indeed adopts isolation, which is the reason to enter into class *I* at a rate of testing). For symptomatic persons, after an average period 1/*γ*_1_*_j_* in class *D*_1_*_j_*, where *γ*_1_*_j_* is the infection rate of pre-diseased persons, pre-diseased persons enter into severe covid-19 class *D*_2_*_j_* (with probability 1 − *m_j_*) or class *Q*_2_*_j_* (with probability *m_j_*), or they are caught by test at a rate *η*_1_*_j_* and enter into class *Q*_1_*_j_*. Persons in class *D*_2_ acquire immunity after period 1/*γ*_2_*_j_*, where *γ*_2_*_j_* is the recovery rate of severe covid-19, and enter into class *I* or die under the disease-induced (additional) mortality rate *α_j_*. Another route of exiting class *D*_2_*_j_* is by treatment, described by the treatment rate *θ_j_*. Class *Q*_1_*_j_* is composed of mild and severe covid-19 persons who came from class *D*_1_*_j_* caught by test, hence they enter into class *D*_2_*_j_* (with the rate (1 − *m_j_*) *γ*_1_*_j_*)or class *I* (with the rate *m_j_ γ*_1_*_j_* + *γ*_2_*_j_* assuming adherence to isolation). Persons in class *Q*_2_*_j_* acquire immunity after period 1/*γ*_3_*_j_*, where *γ*_3_*_j_* is the recovery rate of mild covid-19, and enter into immune class *I*. Another route of exit from class *Q*_2_*_j_* are being caught by a test at a rate *η*_2_*_j_* and enter into class *I* (assumption of adherence to isolation), or enter to class *Q*_3_*_y_* convinced by an education campaign at a rate 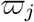, which is temporary, hence *ξ_j_* is the rate of abandonment of protective measures [20].

The initial conditions (*t* = 0) supplied to the system of equations (3), (4), and (5) are, for *j* = *y*, *o*,

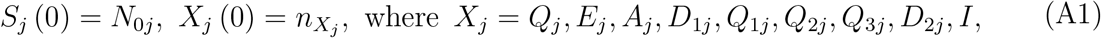

and 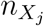 is a non-negative number. For instance, 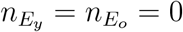 means that there is not any exposed person (young and elder) at the beginning of the epidemic. They are, for young and elder subpopulation,

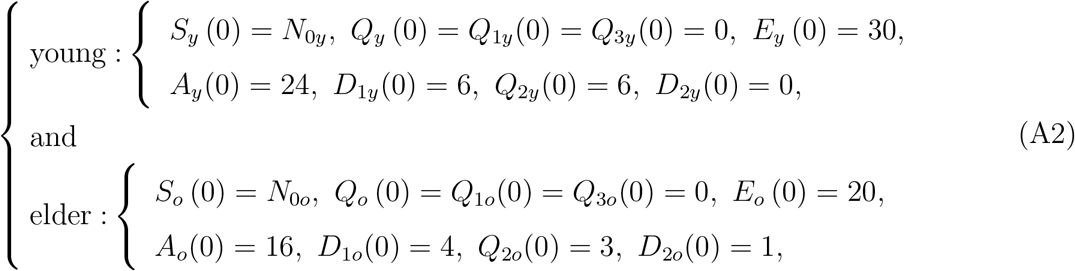

plus *I*(0) = 0, where the initial simulation time *t* = 0 corresponds to the calendar time February 26 when the first case was confirmed in São Paulo State (see [4] [3]). The numbers of young and elder persons are *N*_0_*_y_* = 37.8 million 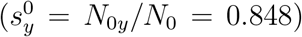 and *N_o_* (0) = *N*_0_*_o_* = 6.8 million 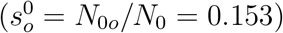, with *N* = *N*_0_*_y_* + *N*_0_*_o_* = 44.6 million.

The isolation implemented at *t* = *τ^is^* (*τ^is^* = 27, when the isolation was implemented on March 24) is described by the boundary conditions

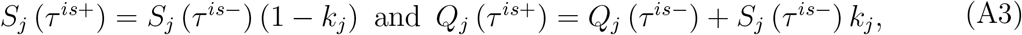

plus

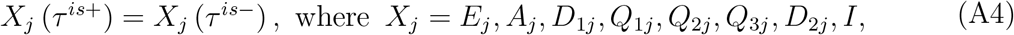

where we have 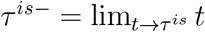 *t* (for *t* < *τ^is^*), and 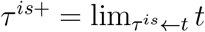 *t* (for *t* > *τ^is^*). If isolation is applied to a completely susceptible population at *t* = 0, there are not any infectious persons, so *S*(0) = *N*_0_. If isolation is done at 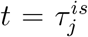 without a screening of persons harboring the virus, then many of the asymptomatic persons could be isolated with susceptible persons.

The boundary conditions supplied to the model at the moments of release *τ_i_*, for *i* = 1,2, ···, m, then 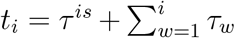, assuming that young and elder subpopulations are released at the same proportion. Hence, the boundary conditions for a series of pulses of release at 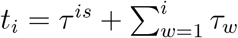, for *i* =1,2, ····, *m*, are

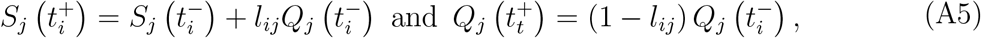

plus

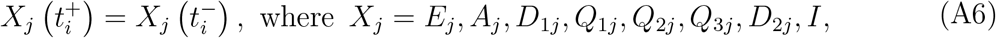

where we have 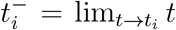 *t* (for *t* < *t_i_*), and 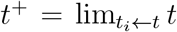 *t* (for *t* > *t_i_*). If *τ_i_* = *τ*, then 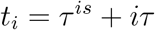.

## Appendix B: The basic reproduction number *R*_0_ – Trivial equilibrium and its stability

The basic reproduction number *R*_0_ is obtained by the analysis of the equilibrium points in the steady-state. However, the non-autonomous and varying population system of equations (2), (4) and (5) does not have steady-state. Disregarding the periods of isolation and release, the system of equations is autonomous (we let *k_j_* = *l_ij_* = 0, *j* = *y*, *o*), but the population *N* varies. For this reason, considering the fractions of individuals in each compartment defined by

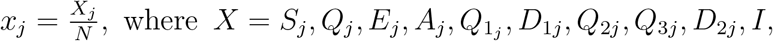

and using equation (6) for *N*, we obtain

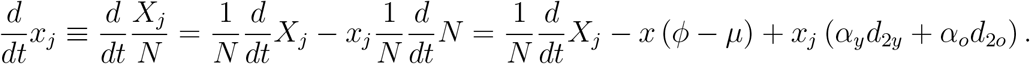

Hence, the system of equations (3), (4) and (5) in terms of fractions become, for susceptible and isolated persons,

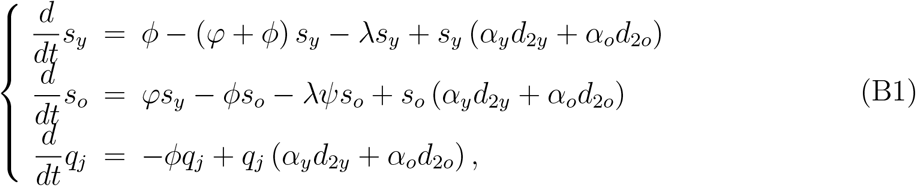

for infected persons,

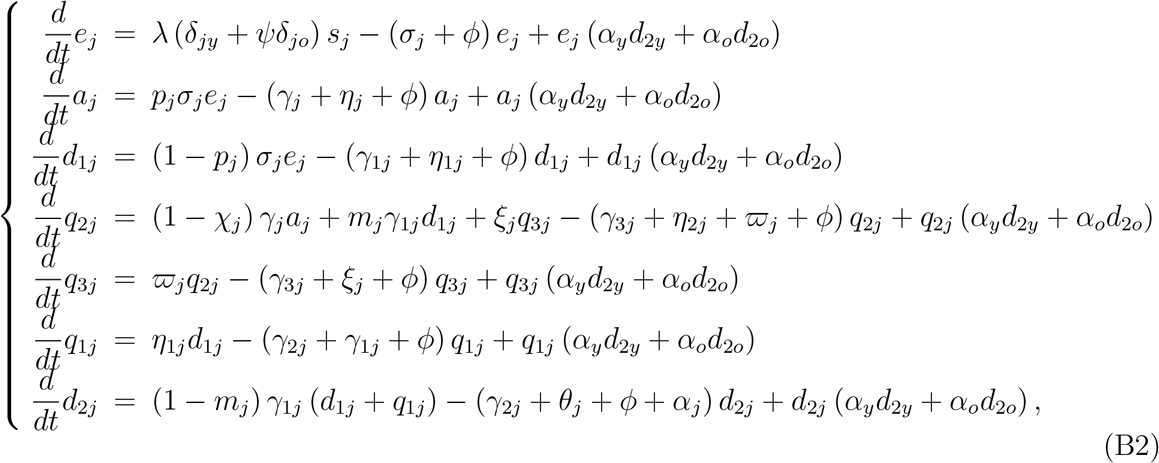

and for immune persons,

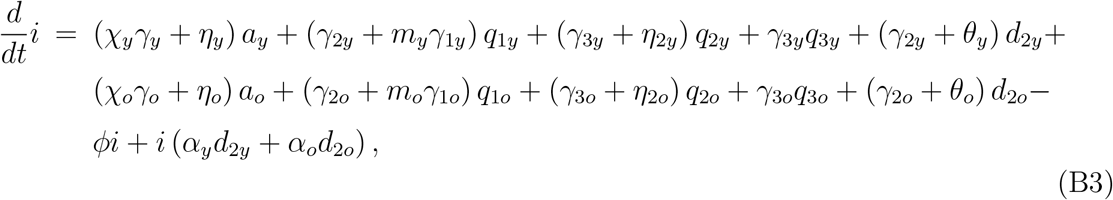

where λ is the force of infection given by equation (1) re-written as

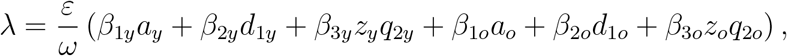

and

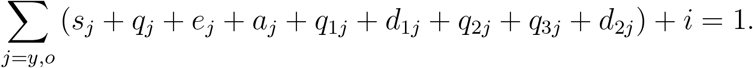

This new system of equation has steady-state, that is, the number of persons in all classes varies with time, however, their fractions attain steady-state (the sum of derivatives of all classes is zero).

The trivial (disease free) equilibrium point *P*^0^ of the system of equations (B1), (B2) and (B3) is given by

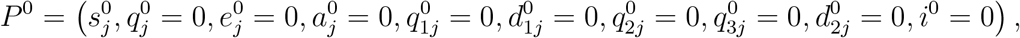

for *j* = *y* and *o*, where

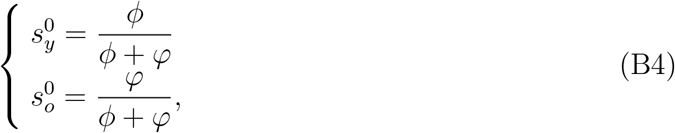

with 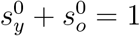.

Let us assess the stability of *P*^0^ by applying the next generation matrix theory considering the vector of variables *x* = (*e_y_*, *a_y_*, *d*_1_*_y_*, *q*_2_*_y_*, *e_o_*, *a_o_*, *d*_1_*_o_*, *q*_2_*_o_*) [21]. We apply method proposed in [22] and proved in [23]. There are control mechanisms, hence we obtain the reduced reproduction number *R_c_* by interventions.

To obtain the reduced reproduction number, diagonal matrix *V* is considered. Hence, the vectors *f* and *υ* are

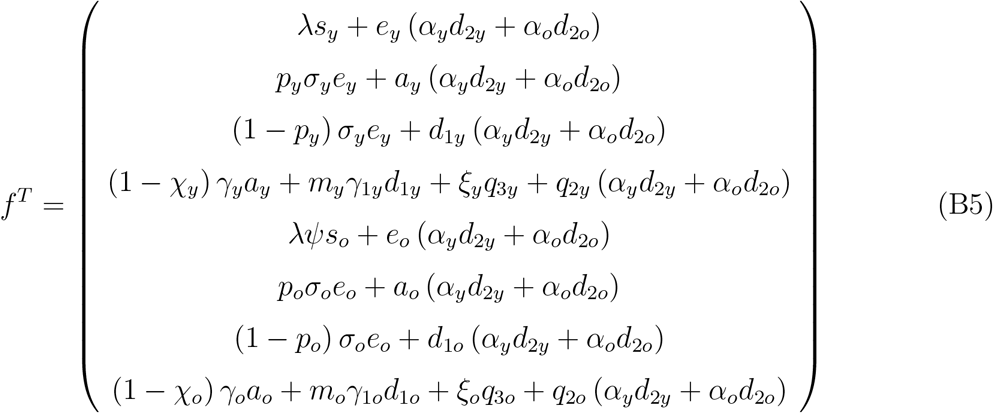

and

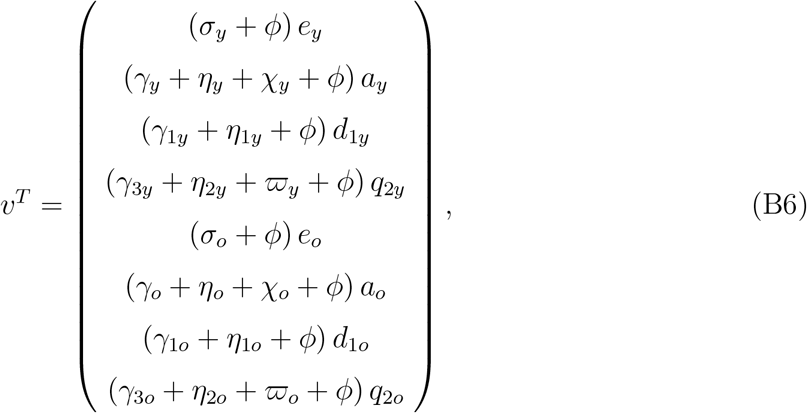

where the superscript *T* stands for the transposition of a matrix, from which we obtain the matrices *F* and *V* (see [21]) evaluated at the trivial equilibrium *P*^0^, which were omitted. The next generation matrix *FV*^−l^ is

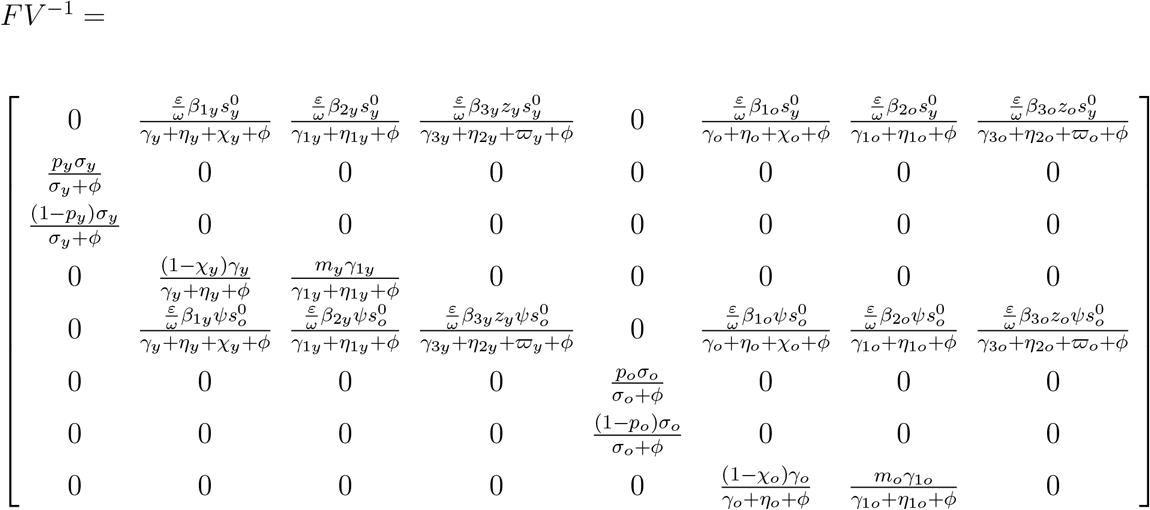

and the characteristic equation corresponding to *FV*^−l^ is

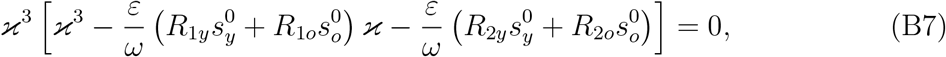

where the partially reduced reproduction numbers *R*_1_*_y_*, *R*_2_*_y_*, *R*_1_*_o_*, and *R*_2_*_o_* are

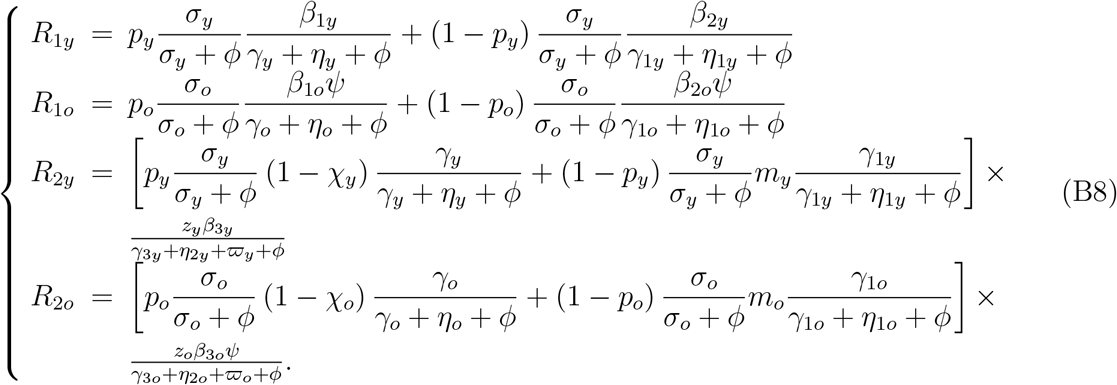

The spectral radius *ρ*(*FV*^−l^) is the biggest solution of a third degree polynomial, which is not easy to evaluate. The procedure proposed in [22] allows us to obtain the threshold *R_c_* as the sum of coefficients of the characteristic equation, where *R_c_* is the reduced reproduction number given by

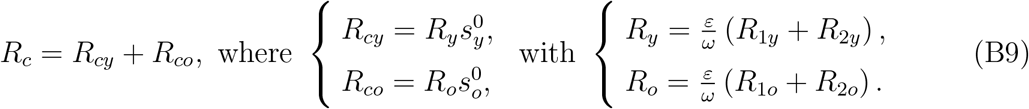

Hence, the trivial equilibrium point *P*^0^ is locally asymptotically stable (LAS) if *R_c_* < 1. The basic reproduction number *R*_0_ is given by

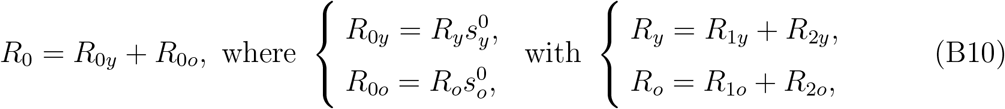

where *R*_1_*_y_*, *R*_2_*_y_*, *R*_1_*_o_*, and *R*_2_*_o_* are obtained from equation (B8) letting *ε* = *ω* = 1 (absence of protective measures), *η_j_* = *η*_1_*_j_* = *η*_2_*_j_* = 0 (absence of test), and *ϖ* = 0 (absence of educational campaign), for *j* = *y*, *o*. All these control actions to decrease the epidemic are absent at the beginning of the epidemic, and the definition of the basic reproduction number is fulfilled.

The basic reproduction number *R*_0_ is the secondary cases produced by one infectious person (could be anyone in one of the classes harboring virus) in a completely susceptible young and elder populations without constraints [6]. Let us understand *R*_1_*_j_* and *R*_2_*_j_*, *j* = *y*, *o*, stressing that the interpretation is the same for both subpopulations, hence we drop out subscript *j*. To facilitate the interpretation, we consider this infectious person in exposed class *E*. This person enters into one of the infectious classes composed of asymptomatic (*A*), pre-diseased (*D*_l_), and a fraction of mild covid-19 (*Q*_2_).

1. *R*_l_ takes into account the transmission by one person in asymptomatic *A* or pre-diseased *D*_l_ class. We interpret for asymptomatic person transmitting (between parentheses, for pre-diseased person) infection. One infectious person survives during the incubation period with probability *σ*/ (*σ* + *ϕ*) and enters into asymptomatic class with probability p (pre-diseased, with 1 − *p*) and generates, during the time 1/ (*γ* + *ϕ*) (pre-diseased, 1/ (*γ*_1_ + *ϕ*)) staying in this class, on average *β*_l_/ (*γ* + *ϕ*) (pre-diseased, *β*_2_/ (*γ*_1_ + *ϕ*)) secondary cases.
2. *R*_2_ takes into account the transmission by a mild covid-19 person. An infectious person has two routes to reach *Q*_2_: passing through *A* or *D*_1_ (this case is given between parentheses). One infectious person survives during the incubation period with probability *σ* / (*σ* + *ϕ*) and enters into asymptomatic (pre-diseased) class with probability *p* (pre-diseased, with 1 − *p*); survives in this class and also is not caught by a test with probability y/ (y + *ϕ*) (pre-diseased, *γ*_1_/ (*γ*_1_ + *ϕ*)) and enters into mild covid-19 class *Q*_2_ with probability 1 − *χ* (pre-diseased, m); and generates, during the time 1/ (*γ*_3_ + *ϕ*) staying in this class, on average z*β*_3_/ (*γ*_3_ + *ϕ*) secondary cases.

Hence, 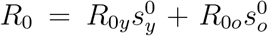 is the overall number of secondary cases generated form one primary case introduced into a completely susceptible young and elder subpopulations. The model parameters are not accurate, and it is expected that *R*_0_ be influenced by the inaccuracy of those values. The variation of *R*_0_ with uncertainties in the parameters can be assessed by the sensitivity analysis [24].

The basic reproduction number *R*_0_ obtained from mathematical modelings provides two useful information: At the beginning of the epidemic (*t* = 0), *R*_0_ gives the magnitude of control efforts, and when epidemic reaches the steady-state (after many waves of the epidemic, that is, t → *∞*), *R*_0_ measures its severity providing the fraction of susceptible individuals, in general, *s*^*^ = 1/*R*_0_ [25]. Between these two extremes, the effective reproduction number *R_ef_* dictates the course of an epidemic, which follows decaying oscillations around *R_ef_* = 1 [26].

Therefore, the importance of a mathematical model is the capability of providing basic and effective reproduction numbers, which provide information about the risk of infection during the epidemic. Our model is complex, but we could obtain *R*_0_, which was not an easy task. However, due to the complexity of the next generation matrix, we could not retrieve the relationsip *s*^*^ = 1/*R*_0_ [25]. The system of equation (B1), (B2) and (B3) has non-trivial (endemic) equilibrium point *P*^*^. One of the steady-state equations of this system allows the definition of the effective reproduction number *R_ef_*. In or model, we have young and elder subpopulations, hence the fraction of susceptible individuals at endemic equilibrium 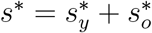 is related to *R*_0_ generically as

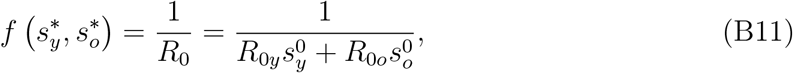

and we define the approximated effective reproduction number *R_ef_* as

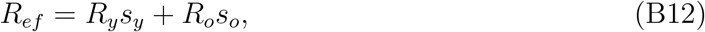

which depends on time.

However, in a simple model, when *z_y_* = *z_o_* = 0 (mild covid-19 cases do not transmit), we have *x* = (*e_y_*, *a_y_*, *d*_1_*_y_*, *e_o_*, *a_o_*, *d*_1_*_o_*). In order to obtain the reduced reproduction number *R_c_*, diagonal matrix *V* is considered. Hence, the vectors *f* and *υ* are

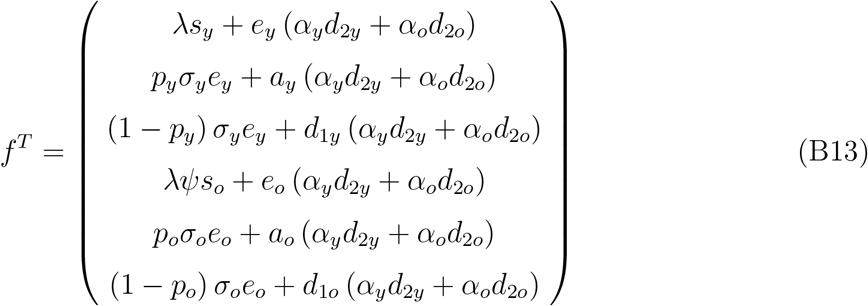

and

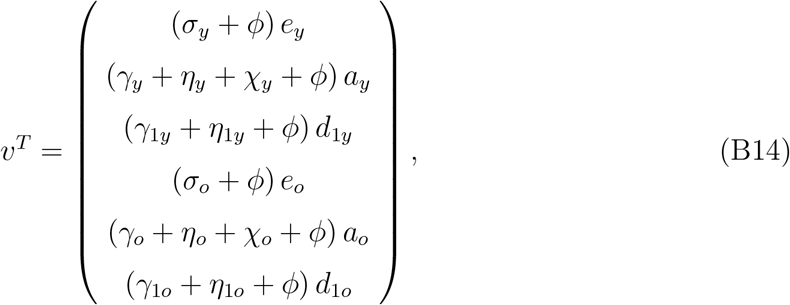

where the superscript *T* stands for the transposition of a matrix, from which we obtain the matrices *F* and *V* (see [21]) evaluated at the trivial equilibrium *P*^0^, which were omitted. The next generation matrix *FV*^−1^ is

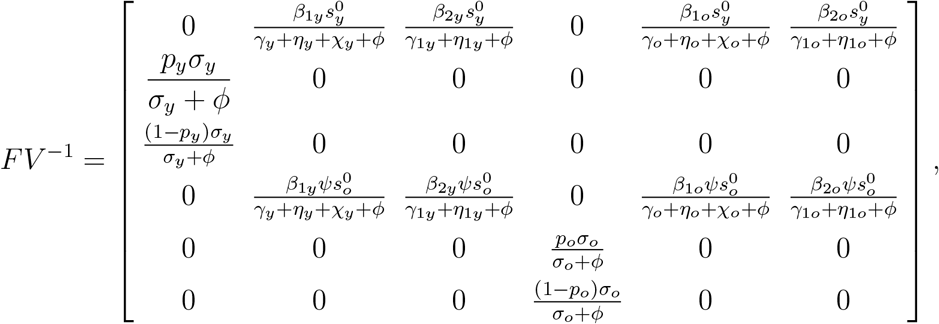

and the characteristic equation corresponding to *FV*^−1^ is

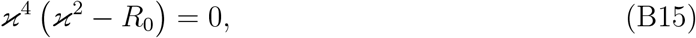

where the basic reproduction number *R_c_* is

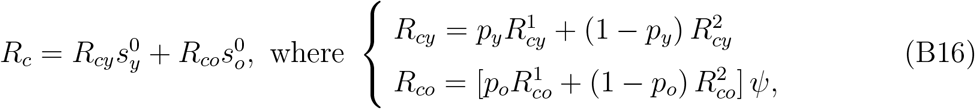

and *R_cy_* and *R_co_* are the basic partial reproduction numbers defined by

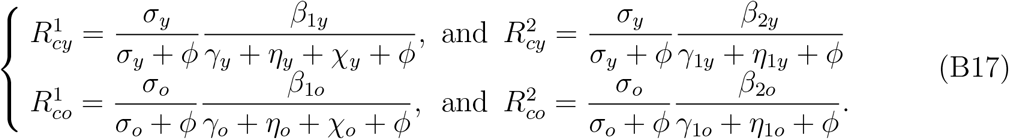

Instead of using the spectral radius 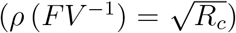, we apply procedure in [22] (the sum of coefficients of characteristic equation), resulting in a threshold *R_c_*. Hence, the trivial equilibrium point *P*^0^ is LAS if *R_c_* < 1.

In order to obtain the fractions of susceptible individuals, *M* must be the simplest (matrix with least number of non-zeros). Hence, the vectors *f* and *υ* are

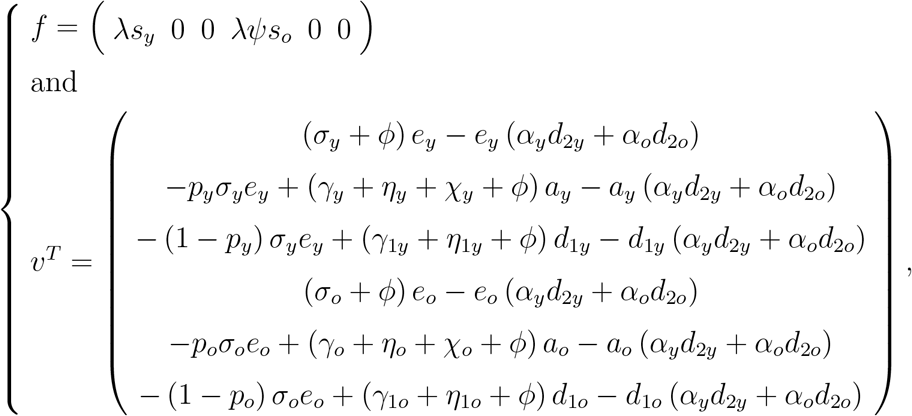

where superscript *T* stands for the transposition of a matrix, from which we obtain the matrices *F* and *V* evaluated at the trivial equilibrium *P*^0^, which were omitted. The next generation matrix *FV*^−1^ is

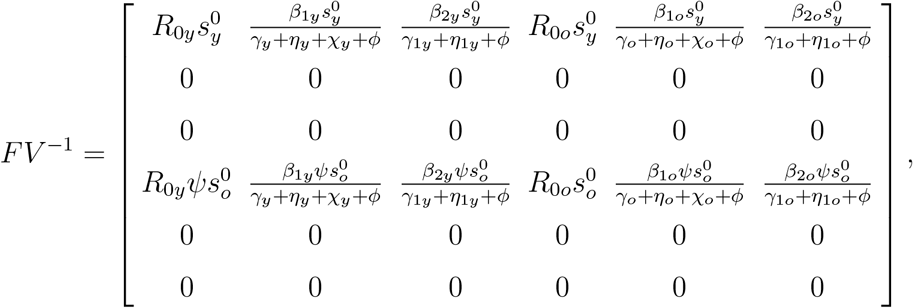

and the characteristic equation corresponding to *FV*^−1^ is

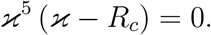

The spectral radius is 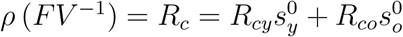 given by equation (B16). Hence, the trivial equilibrium point *P*^0^ is LAS if *ρ* < 1.

Both procedures resulted in the same threshold, hence, according to [25], the inverse of the reduced reproduction number *R_c_* given by equation (B16) is a function of the fraction of susceptible individuals at endemic equilibrium *s*^*^ through

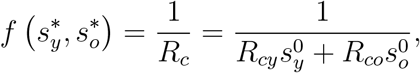

where 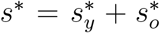 (see [27] [25]). For this reason, the effective reproduction number *R_ef_* [15], which varies with time, can not be defined neither by *R_ef_* = *R*_0_ (*s_y_* + *s_o_*), nor *R_ef_* = *R*_0_*_y_s_y_* + *R*_0_*_o_s_o_*. The function *f*(*ϰ*) is determined by calculating the coordinates of the non-trivial equilibrium point *P*^*^. For instance, for dengue transmission model, 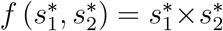, where 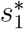 and 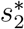 are the fractions at equilibrium of, respectively, humans and mosquitoes [27]. For tuberculosis model considering drug-sensitive and resistant strains, there is not *f*(*ϰ*), but *s*^*^ is solution of a second degree polynomial [25].

The global stability follows method proposed in [28]. Let the vector of variables be x = (*e_y_*, *a_y_*, *d*_1_*_y_*, *e_o_*, *a_o_*, *d*_1_*_o_*), vectors f and v, by equations (B13) and (B14), and matrices *F* and *V* evaluated from f and v at trivial equilibrium *P*^0^ (omitted here). Vector *g*, constructed

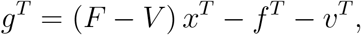

results in

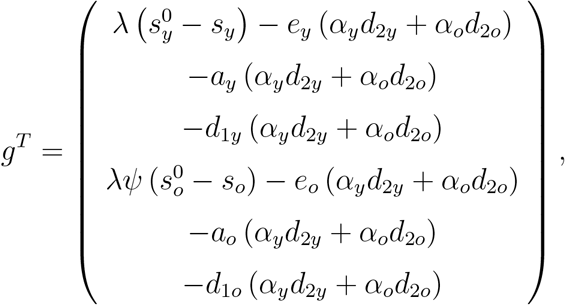

and *g^T^* ≥ 0 if 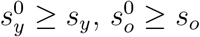 and *α_y_* = *α_o_* =0.

Let *υ_l_* = (*z*_1_, *z*_2_, *z*_3_, *z*_4_, *z*_5_, *z*_6_) be the left eigenvector satisfying *υ_l_V*^−1^ = *F* = *ρυ_l_*, where 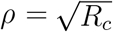, and

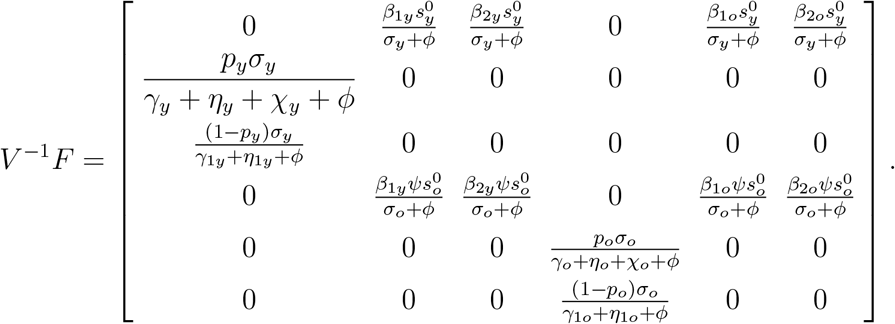

This vector is

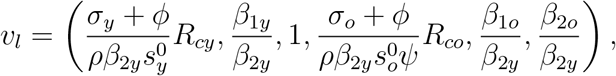

and Lyapunov function *L*, constructed as *L* = *υ_l_V*^−1^*x^T^*, is

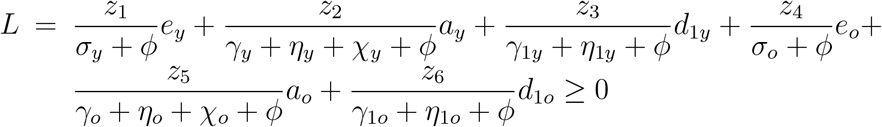

always and

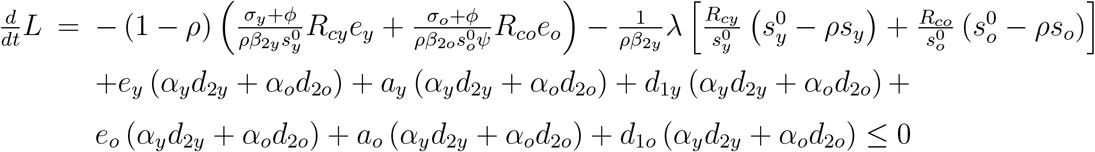

only if *ρ* ≤ 1, 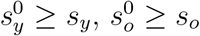 and *α_y_* = *α_o_* = 0.

Hence, the method proposed in [28] is valid only for *α_y_* = *α_o_* = 0 in which case *P*^0^ is globally stable if 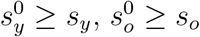 and 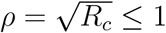.

## Appendix C: Estimation method

To estimate the transmission rates, we assume that all rates in young persons are equal, as well as in elder persons, letting

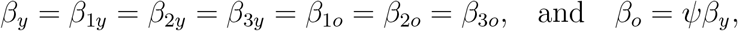

and the forces of infection are *λ_y_* = (*A_y_* + *D*_1_*_y_* + *z_y_Q*_2_*_y_* + *A_o_* + *D*_1_*_o_* + *z_o_Q*_2_*_o_*) *β_y_*/*N* and *λ_o_* = *ψλ*, from equation (1). To estimate the proportion in isolation, we let *k* = *k_y_* = *k_o_*. The model parameters are estimated using data collection of accumulated severe covid-19 cases Ω*^ob^* and deaths П*^ob^* from São Paulo State [8].

To evaluate the parameter *par*, where *par* stands for *β_y_*, *β_o_* = *ψβ_y_*, *k*, *ε* and *ω*, we calculate

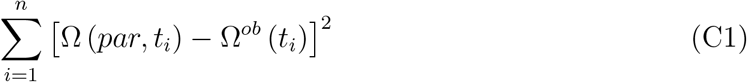

by varying the parameter *par*, and we choose the lower sum of the squared distances between the curve and data. The accumulated covid-19 cases Ω is given by equation (8).

To estimate the mortality rates *α_y_* and *α_o_*, the let *α_y_* = Γ*α_o_*, where Γ is provided by the ratio of deaths occurring in young and elder subpopulations. We evaluate

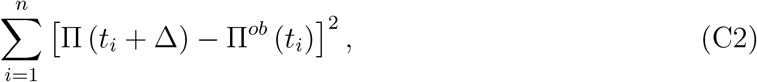

by varying the parameter *α_o_*. The accumulated deaths due to covid-19 П is given by equation (10), with П (0) = 0. Notice that the time of registration *t_i_* of deaths must be related to the deaths of new cases Δ times ago, that is, *D*_2_(*t_i_* − Δ). We use Δ = 15 *days* obtained by analyzing the data from São Paulo State.

Instead of using the system of equations to evaluate the fatality rates *α_y_* and *α_o_* and the recovery rates of severe covid-19 *γ*_2_*_y_* and *γ*_2_*_o_*, we estimate the fatality proportions and the progression rates. The proportions of deaths due to hospitalized covid-19 cases are estimated from data collected from São Paulo State using equations (20), (21), and (22), and minimizing the sum

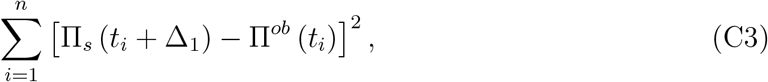

where П_s_ = П_1_ + П_2_ + П_3_ is the sum of all deaths in hospitals.

As we have pointed out about additional mortality rates, in the estimation of fatality proportions, we must consider that the time of registering new cases and deaths is delayed by Δ_1_ days, that is, П_1y_ (*t* + Δ_1_) = *α*_1_*_yς_*_1y_*B*_1_*_y_*(*t*), for instance. The same argument is valid for the number of cured persons, that is, *C_y_*(*t* + Δ _2_) corresponds to *B*_1_*_y_*(*t*) occurred Δ _2_ days ago, for instance.

Siddiqi and Mehra [2] and Pericàs et al. [7] observed three stages in the covid-19 disease states: (I) early infection, (II) pulmonary phase, and (III) hyper inflammation phase. They also observed that complications appeared around days 10-12 after initial symptoms, leading to the cytokine storm. Hence, we let for the period in each one of the three phases 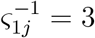, 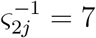 and 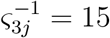, *j* = *y*, *o* (all in days).

At the beginning of epidemics, severe CoViD-19 cases (*D*_2_) were confirmed during hospital care, hence we use *h_y_* = 1.0 and *h_o_* = 1.0. In Wuhan, China [29], 81% of infections did not need hospital care, 14% were severe (developing severe diseases including pneumonia and shortness of breath) and 4.7% were critical (respiratory failure, septic shock, and multi-organ failure). From 19%, we use *h*_1_ = 0.19, which is the proportion of hospitalized persons. However, we use a higher value for elder and lower value for young persons, *h*_1_*_o_* = 0.3 and *h*_1_*_y_* = 0.15. For the ratio hospital:ICUs/intubated, we use approximately 14% and 4.7%, resulting in 3:1, and *h*_2_ = 1/4 = 0.25, which is the proportion of ICUs/intubated care. However, we use higher values for elder and young persons, *h*_2_*_o_* = 0.6 and *h*_2_*_y_* = 0.35. We assume a very small mortality rate in viraemia (phase 1), letting *α*_1_*_y_* = 0.0002 and *α*_1_*_o_* = 0.0006. Patients with inflammatory response in hospital care (phase 2), we let *α*_2_*_y_* = 0.01 and *α*_2_*_o_* = 0.06. Finally, we assume that proportions not surviving in ICUs/intubated with Cytokine storm (phase 3) are *α*_3_*_y_* = 0.45 and *α*_3_*_o_* = 0.65.

